# Proteo-genomics of soluble TREM2 in cerebrospinal fluid provides novel insights and identifies novel modulators for Alzheimer’s disease

**DOI:** 10.1101/2023.06.14.23291409

**Authors:** Lihua Wang, Niko-Petteri Nykänen, Daniel Western, Priyanka Gorijala, Jigyasha Timsina, Fuhai Li, Zhaohua Wang, Muhammad Ali, Chengran Yang, Marta Marquié, Mercè Boada, Ignacio Alvarez, Miquel Aguilar, Pau Pastor, Agustín Ruiz, Raquel Puerta, Adelina Orellana, Jarod Rutledge, Hamilton Oh, Michael D Greicius, Yann Le Guen, Richard J. Perrin, Tony Wyss-Coray, Angela Jefferson, Timothy J. Hohman, Neill Graff-Radford, Hiroshi Mori, Alison Goate, Johannes Levin, Yun Ju Sung, Carlos Cruchaga

**Author notes:** ^17^Lead contact (Carlos Cruchaga). These authors contributed equally (Lihua Wang, Niko-Petteri Nykänen). ^¶^ These authors contributed equally. Corresponding author information: Carlos Cruchaga, PhD Washington University, School of Medicine 425 S. Euclid Ave. BJC Institute of Health. Box 8134 St. Louis, MO 63110.

## Abstract

Triggering receptor expressed on myeloid cells 2 (TREM2) plays a critical role in microglial activation, survival, and apoptosis, as well as in Alzheimer’s disease (AD) pathogenesis. We previously reported the *MS4A* locus as a key modulator for soluble TREM2 (sTREM2) in cerebrospinal fluid (CSF). To identify additional novel genetic modifiers of sTREM2, we performed the largest genome-wide association study (GWAS) and identified four loci for CSF sTREM2 in 3350 individuals of European ancestry. Through multi-ethnic fine mapping, we identified two independent missense variants (p.M178V in *MS4A4A* and p.A112T in *MS4A6A*) that drive the association in *MS4A* locus and showed an epistatic effect for sTREM2 levels and AD risk. The novel *TREM2* locus on chr 6 contains two rare missense variants (rs75932628 p.R47H, P=7.16×10^-19^; rs142232675 p.D87N, P=2.71×10^-10^) associated with sTREM2 and AD risk. The third novel locus in the *TGFBR2* and *RBMS3* gene region (rs73823326, P=3.86×10^-9^) included a regulatory variant with a microglia-specific chromatin loop for the promoter of *TGFBR2*. Using cell-based assays we functionally validated that overexpression of *TGFBR2* increased sTREM2 and silencing reduced sTREM2, whereas modulating *RBMS3* did not. The last novel locus *NECTIN2* on chr 19 (rs11666329, P=2.52×10^-8^) was independent of *APOE* genotype and colocalized with cis-eQTL of *NECTIN2* in the brain cortex and cis-pQTL of NECTIN2 in CSF. To our knowledge, this is the largest study to date aimed at identifying genetic modifiers of CSF sTREM2. This study provided novel insights into the *MS4A* and *TREM2* loci, two well-known AD risk genes, and identified *TGFBR2* and *NECTIN2* as additional modulators involved in TREM2 biology.

## Background

Alzheimer’s disease (AD) is a progressive neurodegenerative disorder, associated with irreversible memory deficits and cognitive decline. Extracellular amyloid-β (Aβ)-containing plaques, intracellular accumulated tau neurofibrillary tangles, and neuroinflammation are the main pathological changes observed in AD patients. Apolipoprotein E (*APOE*) on chromosome 19 is the strongest genetic risk factor for late-onset AD [1]. In 2013, two groups independently identified the rare *TREM2-*p.R47H that increased AD risk almost threefold, similar to the *APOE* ε4 allele [2, 3]. Additional rare variants in *TREM2* including p.R62H, p.T96K, p.D87N, p.H157Y, p.R98W, p.T66M, p.Y38C and p.Q33X were subsequently identified to be associated with AD [3-6] In addition, the latest genome-wide association studies (GWAS) including low-frequency variants identified the *TREM2* locus for AD [7, 8].

TREM2 is an innate immune response receptor and type I transmembrane protein, highly expressed in microglia [9]. TREM2 plays important roles in microglia activation, survival, migration, and phagocytosis [10]. Microglia have been implicated in AD via phagocytosing dead cells, eliminating Aβ plaques, and pruning synaptic connection [11-13]. Therefore, dysregulated microglia function in the brain due to *TREM2* risk variants may increase AD risk. Failure of microglia migration to Aβ plaques augmented insoluble Aβ_40_ and Aβ_42_ accumulation and increased neural dystrophy in Trem2^−/−^ 5XFAD mice [14]. These pathological alterations and impaired cognitive function were rescued in human *TREM2* bacterial artificial chromosome (BAC) transgenic mice [15]. In contrast, TREM2 is detrimental to tau pathology and *Trem2* deficiency protects against neurodegeneration in PS19 human tau transgenic mice [16]. These studies together demonstrate that TREM2 plays an important, but complex, role in AD pathology.

A soluble form of TREM2 (sTREM2) in cerebrospinal fluid (CSF) has emerged as an important biomarker for AD progression and pathogenesis. The full-length TREM2 protein consists of an extracellular ectodomain, a transmembrane domain, and an intracellular domain [17]. Among the three major *TREM2* transcripts found in human brains [5, 18], an alternative spliced transcript (ENST00000338469) excludes exon 4, which encodes the transmembrane domain, and produces sTREM2 [5]. In addition, sTREM2 can be produced by proteases including ADAM17, ADAM10, or γ-secretases [19]. We and other groups have shown that CSF sTREM2 is elevated in AD [20-22]. In autosomal dominant AD, the changes in CSF sTREM2 occur 5 years before the expected onset of AD [23]. CSF sTREM2 is positively correlated with CSF tau and phosphorylated tau (P-tau) at threonine 181, but not with Aβ42, indicating that sTREM2 is associated with neurodegeneration after amyloid accumulation [21, 22, 24]. Higher sTREM2 in CSF is also shown to be associated with slower cognitive decline in AD [25, 26]. However, other proteins that are part of the TREM2 and sTREM2 pathways, and the downstream mechanism by which these proteins lead to AD are still unknown.

We previously performed a GWAS for CSF sTREM2 and identified the *MS4A* locus which included *MS4A4A* and *MS4A6A* among others on chromosome 11 as a key modulator of CSF sTREM2 levels [27, 28]. We demonstrated that MS4A4A and TREM2 colocalized to lipid rafts at the plasma membrane and that *MS4A4A* modified sTREM2 in a dose dependent manner. However, in that study we were not able to identify the functional variant driving the association in the *MS4A* locus, nor to determine whether there were any additional functional genes in this or other loci modifying sTREM2 levels.

In this study, we performed a GWAS of CSF sTREM2 levels and identified four loci in a large cohort that included 3,350 non-Hispanic European ancestry (EURs) individuals. To further pinpoint the functional variants and nominate the functional gene underpinning the identified loci, we then performed post-GWAS analyses including multi-ethnic fine mapping with 250 non-European individuals (non-EURs), stepwise conditional analyses, colocalization analyses, annotation with brain cell-type specific enhancer-promoter interaction*, in vitro* functional validation, and Mendelian randomization analysis.

## Results

### GWAS analyses identified four loci associated with CSF sTREM2 levels

To identify novel genetic variants that modify CSF sTREM2 protein levels, we performed GWAS of CSF sTREM2 in 3,350 unrelated EURs individuals from eight cohorts (Fig. S1, Table 1, and Fig. 1A). There was no evidence for genomic inflation (λ_GC_=1.004 in EURs; Fig. S2; λ_GC_=1.015 in non-EURs; Fig. S3). A total of four loci reached genome-wide significance (P < 5×10^-8^; Fig. 1B, Fig. 1C and Table 2). The most significant locus was tagged by a common variant (rs72918674, P=4.29×10^-62^) with minor allele frequency (MAF) of 0.398 on chromosome 11 within the *MS4A* gene region that we previously reported [27]. The second significant locus was tagged by a rare variant (rs12664332, MAF=0.006, β=-1.39, P=2.25×10^-20^) on chromosome 6 in the *TREM2* gene region. In addition, we observed a third locus tagged by a low frequency intergenic variant (rs73823326, MAF=0.06, β=-0.282, P=3.86×10^-9^) on chromosome 3 between the *RBMS3* and *TGFBR2* genes. Finally, we observed a common variant (rs11666329, MAF=0.496, β=0.126, P=2.52×10^-8^) on chromosome 19 within the *APOE* region that also passed the genome-wide threshold. With the exception of the *MS4A* locus, all three other loci were identified for the first time to be associated with sTREM2 levels (Fig. 1B).

**Fig. 1.**
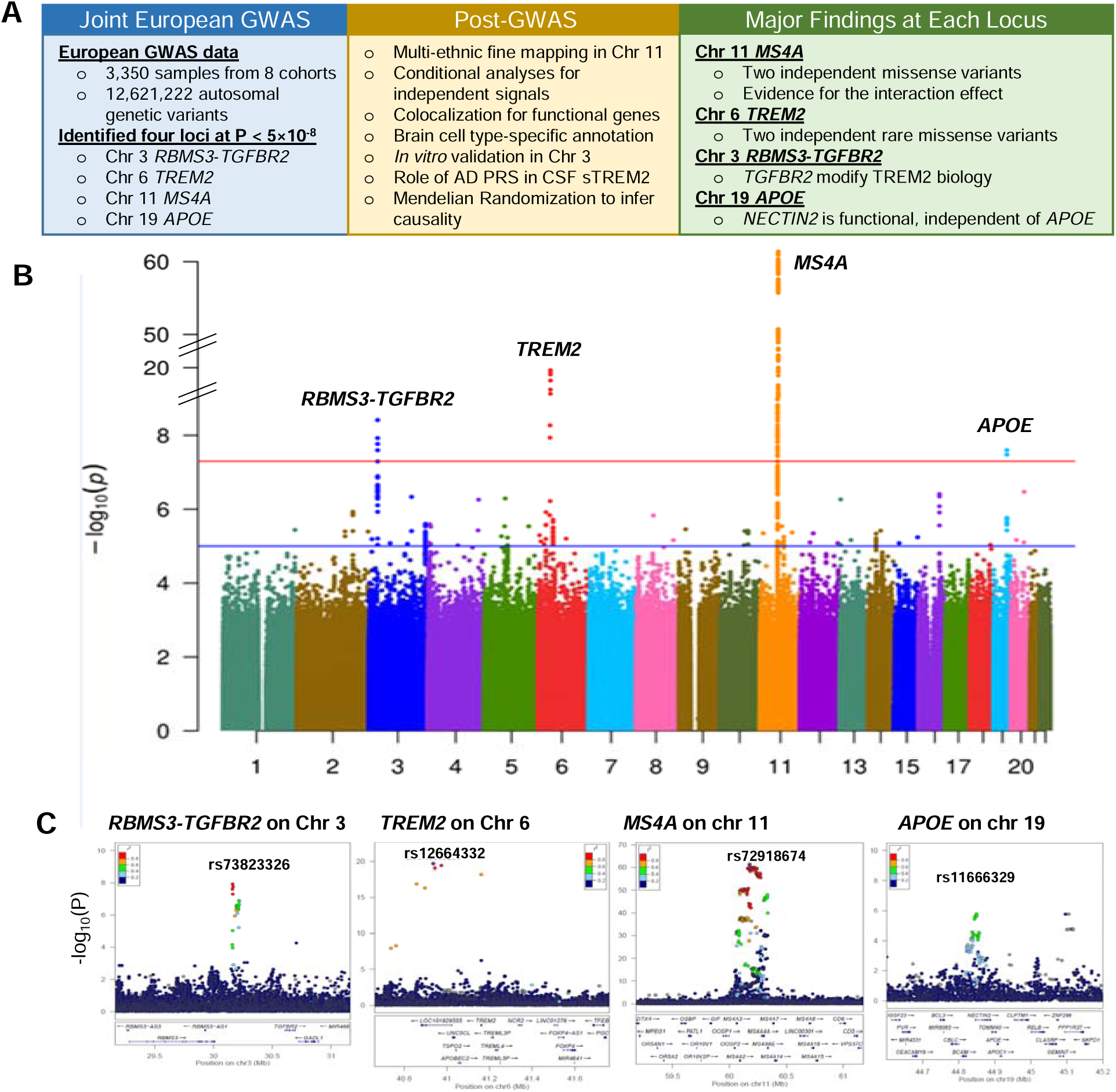
Workflow of proteo-genomics of soluble TREM2 (sTREM2) in cerebrospinal fluid (CSF) and association results using 3350 EUR samples. **A)** Our study included two stages of analyses: first stage GWAS analyses using 3350 European samples from eight cohorts and 12,621,222 autosomal genotypic variants, and second stage multi-ethnic fine mapping using 250 non-European (non-EUR) samples from eight cohorts and 8,909,120 autosomal genotypic variants. CSF sTREM2 was measured by SomaScan or MSD. In the first-stage GWAS analyses, using an additive linear model adjusting for age at CSF draw, sex, genotype platform/cohorts, and 10 PCs, we identified 4 loci associated with CSF sTREM2 levels: chromosome 3 RBMS3-TGFBR2 (novel), chromosome 6 TREM2 (novel), chromosome 11 MS4A (known), and chromosome 19 APOE (novel) as shown in Manhantan plot and locus zoom plots. For these 4 loci, we then conducted post-GWAS analyses. First, we used multi-ethnic fine mapping to detect the true causal variants underlying each locus. For each of the four loci, we then performed stepwise conditional analyses to identify the independent genotypic variants. To identify the functional genes underlying three novel loci, we performed colocalization analyses of each locus with the AD GWAS, GTEx eQTL, and MetaBrain eQTL. The regulatory role of these loci were annotated with the brain cell type-specific enhancer-promoter interaction map. For chromosome 3 RBMS3-TGFBR2 locus, in vitro functional validation using overexpression of TGFBR2 and RBMS3 in human primary macrophages was conducted. The overall genetic architecture overlapped between AD PRS and CSF sTREM2 was estimated using multivariate linear regression. Finally to determine whether CSF sTREM2 is causal for AD, two-sample Mendelian randomization was analyzed using CSF sTREM2 GWAS as exposure and the latest AD GWAS as outcome. **B)** Manhantan plots of GWAS for cerebrospinal fluid (CSF) soluble triggering receptor expressed on myeloid cells 2 (sTREM2) in European individuals (EURs). P values are two-sided raw P values estimated from a linear additive model. The blue solid horizontal line denotes the genome-wide significance level (P = 5 × 10^-8^), and the red solid horizontal line represents the suggestive significance level (P = 1 × 10^-6^). X-axis depicts genomic coordinates by chromosome number and y-axis denotes the negative log10-transformed P value for each genetic variant. **C)** LocusZoom plot of GWAS of CSF sTREM2 at chromosome 3, 6, 11, and 19. The X-axis depicts genomic coordinates and the y-axis denotes the negative log10-transformed P value for each genetic variant.

**Table 1.** Characteristics of sample by cohorts for EURs participants.

| Cohort | N | Age (mean ± SD) | Female (%) | APOE ε4+ (%) | Cases (%) | sTREM2 [5635-66 |
| --- | --- | --- | --- | --- | --- | --- |
| **Knight ADRC** | 797 | 71.36 ± 8.65 | 425 (53.32 ) | 314 (39.4) | 178 (22.33) | 0.36 ± 0.93<sup>7k</sup> |
| **ADNI** | 676 | 73.75 ± 7.47 | 283 (41.86 ) | 342 (50.59) | 512 (75.74) | -0.3 ± 0.94<sup>7k</sup> |
| **ACE** | 435 | 71.95 ± 8.28 | 257 (59.08 ) | 156 (35.86) | 238 (54.71) | 0.08 ± 0.94<sup>7k</sup> |
| **Barcelona-1** | 187 | 68.82 ± 7.38 | 90 (48.13 ) | 77 (41.18) | 59 (31.55) | 0.07 ± 1.08<sup>7k</sup> |
| **DIAN** | 172 | 39.89 ± 10.64 | 92 (53.49 ) | 48 (27.91) | 118 (68.6) | -0.7 ± 0.92<sup>7k</sup> |
| **PPMI** | 779 | 61.12 ± 9.33 | 330 (42.36 ) | 174 (22.34) | 460 (59.05) | 0.01 ± 0.99<sup>5k</sup> |
| **VMAP** | 135 | 72.39 ± 6.42 | 42 (31.11 ) | 49 (36.3) | 51 (37.78) | 0.03 ± 0.96<sup>MSD</sup> |
| **Stanford ADRC** | 169 | 69.73 ± 6.66 | 97 (57.4 ) | 68 (40.24) | 46 (27.22) | 0.12 ± 1.00<sup>5k</sup> |

Knight ADRC = Charles F. and Joanne Knight Alzheimer Disease Research Center
ADNI = Alzheimer's Disease Neuroimaging Initiative
ACE = ACE Alzheimer Center Barcelona
Barcelona-1 = Longitudinal observational study from the Memory and Disorder unit at the University Hospital Mutua de Terrassa, Terrassa, Barcelona, Spain.
DIAN = Dominantly Inherited Alzheimer Network
PPMI = Parkinson's Progression Markers Initiative
VMAP=Vanderbilt Memory and Aging Project
Stanford ADRC = Stanford Alzheimer Disease Research Center
Age is age at Lumbar puncture (LP). Values denote years in mean ± SD.

**Table 2.**
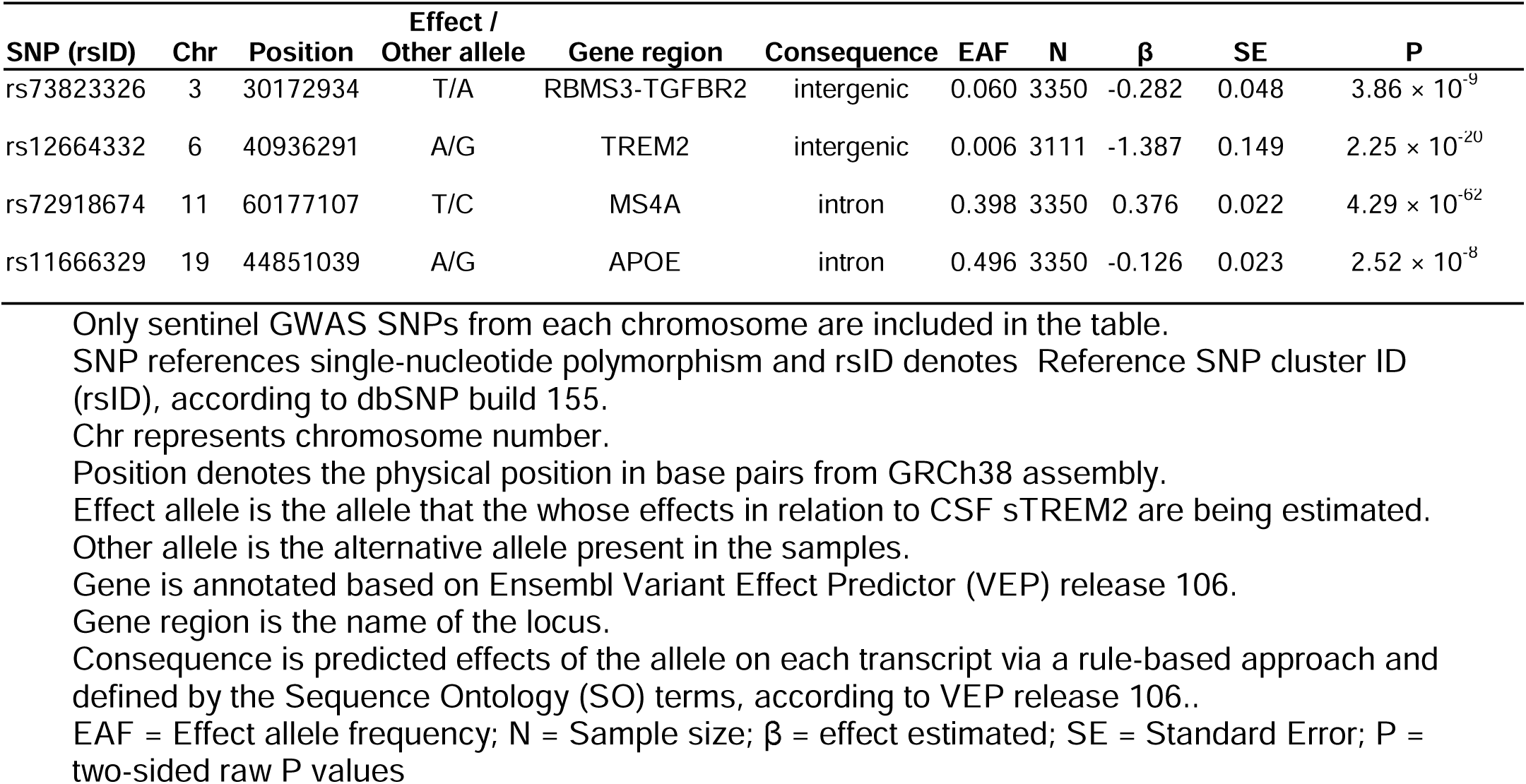
Summary of association results in the European individuals (EUR)

As our datasets were enriched for Alzheimer disease cases, we wanted to determine if any of the associations were influenced by disease status. To do this, we performed five different sensitivity analyses of the sentinel variants: 1) Association analyses adjusted for AD status (n=1,972); 2) Association analyses adjusted for biomarker positivity based on the ATN classification (n=1,600); 3) Association analyses adjusted for Clinical Dementia Rating (CDR; n=1,639); 4) Association analyses only using biomarker negative (A-T-) individuals (n=841); 5) Association analyses only using biomarker positive (A+T+) individuals (n=759). We found a very high correlation of the effect sizes in each analyses (Fig. S4, R=1, p< 5.3×10^-4^). In addition, the effect sizes of these five models were not significantly different to our main model (Table S1). Our findings indicate that the genetic regulation of CSF sTREM2 was not affected by clinical or biomarker status.

### Tissue Specificity of four Identified genetic loci

In order to determine whether four identified genetic loci are specific for CSF, we examined the association of these four loci with plasma sTREM2 based on 35,559 Icelanders (Table S2). The loci at chromosome 6 at the *TREM2* locus, in cis, and chromosome 11 at *MS4A* locus, showed highly significant association (P < 1.5×10^-250^; Table S2) in plasma, indicating shared genetic regulation between CSF and plasma. Notably, our two new signals, chromosome 3 (*TGFBR*2/*RBMS3*) and chromosome 19 (*NECTIN2/APOE*), did not showed any association with plasma sTREM2 (P>0.17), suggesting these are CSF specific signals and reinforming the notion that it is important to study relevant tissues other than plasma for Alzheimer’s disease.

### Multi-ethnic fine mapping identified two independent functional variants and genes in *MS4A* modifying sTREM2 and AD risk

The *MS4A* locus on chromosome 11 showed the most significant association for CSF sTREM2 levels (Fig. 1B). This locus included 488 genetic variants reaching genome-wide significance (all with P < 5×10^-8^; Table S3). The sentinel variant was a common variant (rs72918674, MAF=0.40, β=0.38, P=4.29×10^-62^) located within an intron of *MS4A6A*.

In order to determine the presence of an additional independent signal in this locus, we performed conditional analysis. After conditioning by the sentinel variant rs72918674, a secondary signal located within an intron of *MS4A4A* (rs10897026, MAF=0.32; β=-0.28, P=2.98×10^-31^ before conditioning; β=-0.19, P=6.38×10^-16^ after conditioning; Fig. 2A) was identified. There were no additional independent signals beyond these two tagged by rs72918674 and rs10897026. The linkage disequilibrium (LD) structure of this region revealed that these two signals belong to two distinct LD blocks (r^2^=0.06 between two index variants; Fig. 2B and Table S3). For these two signals, all eight cohorts contributed consistently to the association, without any evidence of heterogeneity (heterogeneity P=0.36 for rs72918674 and P=0.78 for rs10897026; Fig. 2C).

**Fig. 2.**
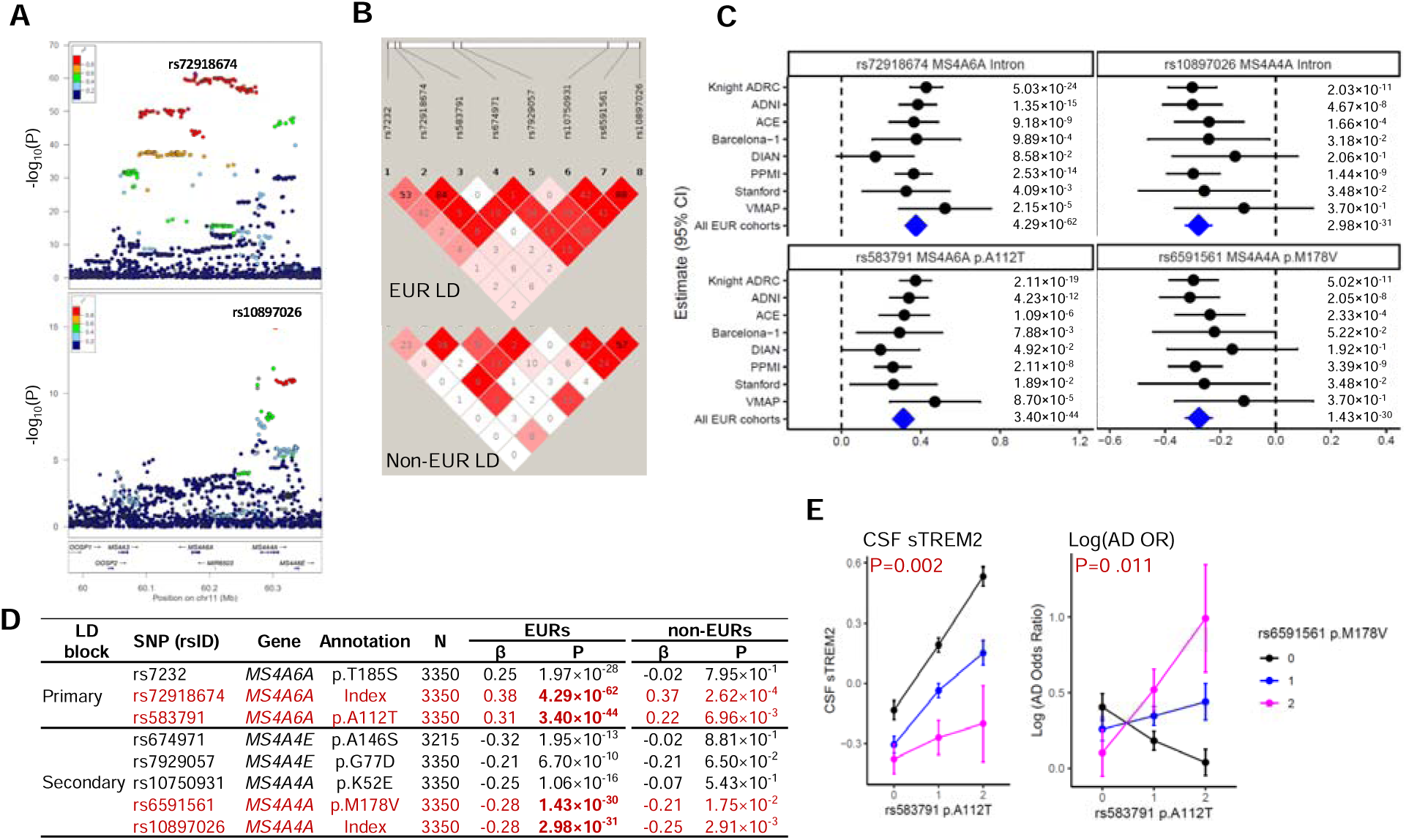
Association results of CSF sTREM2 at chromosome 11. **A)** LocusZoom plots at chromosome 11 in European ancestry (EURs) for the sentinel SNP rs72918674 and the secondary signal rs10897026 conditioning on the sentinel SNP. X-axis depicts genomic coordinates at chromosome 11 and y-axis denotes the negative log10-transformed P value for each genetic variant. **B)** Linkage disequilibrium (LD) heatmap of chromosome 11 SNPs in EURs and non-EURs. **C)** Forest plots of effect size estimates by cohort for rs72918674 (MS4A6A intron), rs10897026 (MS4A4A intron), rs583791 (MS4A6A, p.A112T) and rs rs6591561 (MS4A4A, p.M178V). Heterogeneity P is 3.69 × 10^-1^ for rs72918674, 4.10 × 10^-1^ for rs583791, 7.79 × 10^-1^ for rs10897026, and 7.92 × 10^-1^ for rs6591561 respectively. **D)** Summary of association results of 2 independent SNPs and 6 missense variants in the *MS4A* gene region from EURs. SNP references single-nucleotide polymorphism and rsID denotes reference SNP cluster ID (rsID), according to dbSNP build 155. Gene is annotated based on Ensembl Variant Effect Predictor (VEP) release 106. Annotation is the definition of identified SNP as Top Hit, Secondary, or amino acid changes. N is the sample size in GWAS of European (EURs) samples. β in EURs is effect estimated in EURs samples. P in EURs is two-sided raw P values in EURs samples. β in non-EURs is effect estimated in non-European (non-EURs) samples. P in non-EURs is two-sided raw P values in non-EURs samples. **E)** Effect of epistasis between rs583791 (MS4A6A, p.A112T) and rs6591561 (MS4A4A, p.M178V) on CSF sTREM2 levels and Log of Alzheimer’s disease (AD) Odds ratio. X-axis is dosage of rs583791 (MS4A6A, p.A112T) coded based on the copy of C allele. Y-axes are Z-score of CSF sTREM2 and Log of AD Odds ratio. The color is based on the dosage of rs6591561 (MS4A4A, p.M178V) coded based on the copy of G allele. The effect allele is T for rs72918674, C for rs583791, G for rs6591561, and C for rs10897026. The association of the first variant rs583791 is much stronger in individuals with CC genotype of the second variant rs6591561, as shown in the steep black line, when compared to that for those with TT genotype.

To identify the most likely functional variant(s), we performed functional annotation of the associated variants. The primary signal, rs72918674, is in LD with two missense variants in *MS4A6A* (rs7232 p.T185S; rs583791 p.A112T; first LD block; Table S3; Fig. S5A; Fig. 2B). The secondary signal, rs1089702, is in LD with two missense variants (rs10750931 p.K52E; rs6591561 p.M178V; second LD block; Table S3; Fig. S5A; Fig. 2B) in *MS4A4A* as well as in LD in two missense variants (rs674971 p.A146S; rs7929057 p.G77D) in *MS4A4E*.

While some of the missense variants may be the causal variants modifying sTREM2 levels, the remaining variants would be in LD with the functional variants. As LD structure varies across populations due to random genetic drift, genetic mutation, and recombination events [29],analyses of other ethnicities can help distinguish the functional variants from those in LD. To achieve this goal of fine-mapping, we performed an association analysis in 250 non-EURs (mostly African American individuals while including some of Asian and Hispanic ancestry; Table S4 and Fig. S1C). In the first LD block that contained the primary signal, one missense variant p.A112T in *MS4A6A* (P=6.96×10^-3^) remained significant at P < 0.05. This missense variant had an LD r^2^ of 0.363 with the primary index variant in the non-EURs population (Table S5 and Fig. 2B). The effect sizes for these variants were consistent across populations (Table S3). In the second LD block that contained the secondary signal, only one missense variant p.M178V in *MS4A4A* (P=1.75×10^-2^) was significant in the non-EURs, with consistent effect size (β=-0.278 in EURs vs. -0.209 in non-EURs). These results suggest that the *MS4A6A* p.A112T and the *MS4A4A* p.M178V variants are the major functional SNPs driving the association in this locus. Results of the remaining variants in this locus were included in Table S3. However, the effects of the variants are in opposite direction, with minor allele of p.A112T being associated with higher CSF sTREM2, and the minor allele of p.M178V being associated with lower CSF sTREM2. In both case, the allele associated with higher CSF sTREM2 levels is associated with lower AD risk.

We hypothesize that these two independent signals in this locus could have a synergistic effect. We therefore performed epistatic analysis for CSF sTREM2 and AD risk by including *MS4A6A* p.A112T, *MS4A4A* p.M178V, and their interaction term in a linear model. We found a significant interaction among these two variants for both CSF sTREM2 levels (P=0.002; Table S6 and Fig. 2E) and AD risk (P=0.011; Table S7 and Fig. 2E). This indicates that these two missense variants are jointly affecting CSF sTREM2 levels as well as AD risk. However, the underlying molecular mechanism for this identified interaction needs further investigation.

We subsequently examined the association of these two functional variants with AD risk and related phenotypes (Table S8). The variant *MS4A6A* p.A112T showed a strong association with AD risk (P=6.98×10^-17^) [8]. In addition, the association in this locus for CSF sTREM2 and AD risk had very strong evidence for colocalization (PP.H4=0.96; Table S9, Fig. S5B). Moderate association (P=4.72×10^-4^) with AD risk was found with the second variant *MS4A4A* p.M178V [30]. For both independent signals, the allele associated with higher CSF sTREM2 levels was associated with lower AD risk (Table S8). In addition to the association with AD risk, this locus is also associated with other AD endophenotypes. Specifically, the alleles associated with higher CSF sTREM2 levels and lower AD risk was also associated with lower (protective) CSF pTau (β=-2.65, P=8.11×10^-3^) [31], significantly later age at onset for AD (β=-0.06, P=3.64×10^-7^; Table S8) [32], and showed suggestive association with lower rate of memory decline (β=-0.07, P=6.48×10^-2^) [33].

### The association with sTREM2 at the *TREM2* locus is driven by two *TREM2* missense variants

The *TREM2* locus on chromosome 6 was the second most significant locus for sTREM2 levels (Fig. 3A). This locus contains nine genetic variants reaching genome-wide significance. Two of these significant variants are missense variants: p.R47H and p.D87N for *TREM2*. The index variant (rs12664332, MAF=0.006, β=-1.39, P=2.25×10^-20^) was in LD (r^2^=0.799) with one missense variant (p.R47H, MAF=0.006, β=-1.349, P=7.16×10^-19^) of *TREM2* (top panel of Fig. 3A) as well as other seven variants in this region (Fig. S6A). The conditional analysis identified an additional independent signal at missense variant (rs142232675, p.D87N, MAF=0.003, β=- 1.843, P=2.71×10^-10^) which is also in the other LD block (bottom panel of Fig. 3A; LD between these two missense variants r^2^=0). The minor alleles of both missense variants were associated with lower CSF sTREM2 levels, with a consistent effect size across six cohorts (Fig. 3B and Fig. 3C). Colocalization analysis with the latest AD GWAS [8] confirmed that this locus is the same as the one for AD risk (PP.H4=1.00; Fig. S6A and S6B). The two missense variants (p.R47H and p.D87N) in *TREM2* were previously identified for AD risk, but not for CSF sTREM2 levels. In addition, a suggestive association was observed for the P.R62H variant, which has also been reported to be associated with AD risk(rs143332484, MAF=0.011, β=-0.55, P=6.02×10^-7^; Fig. 3B and Fig. 3C). This missense variant is not in LD with either rs142232675 p.D87N (r^2^=0) or rs75932628 p.R47H (r^2^=0.0001). Our previous study [27] included 800 samples and was unable to identify this locus due to the low frequency of these variants.

**Fig. 3.**
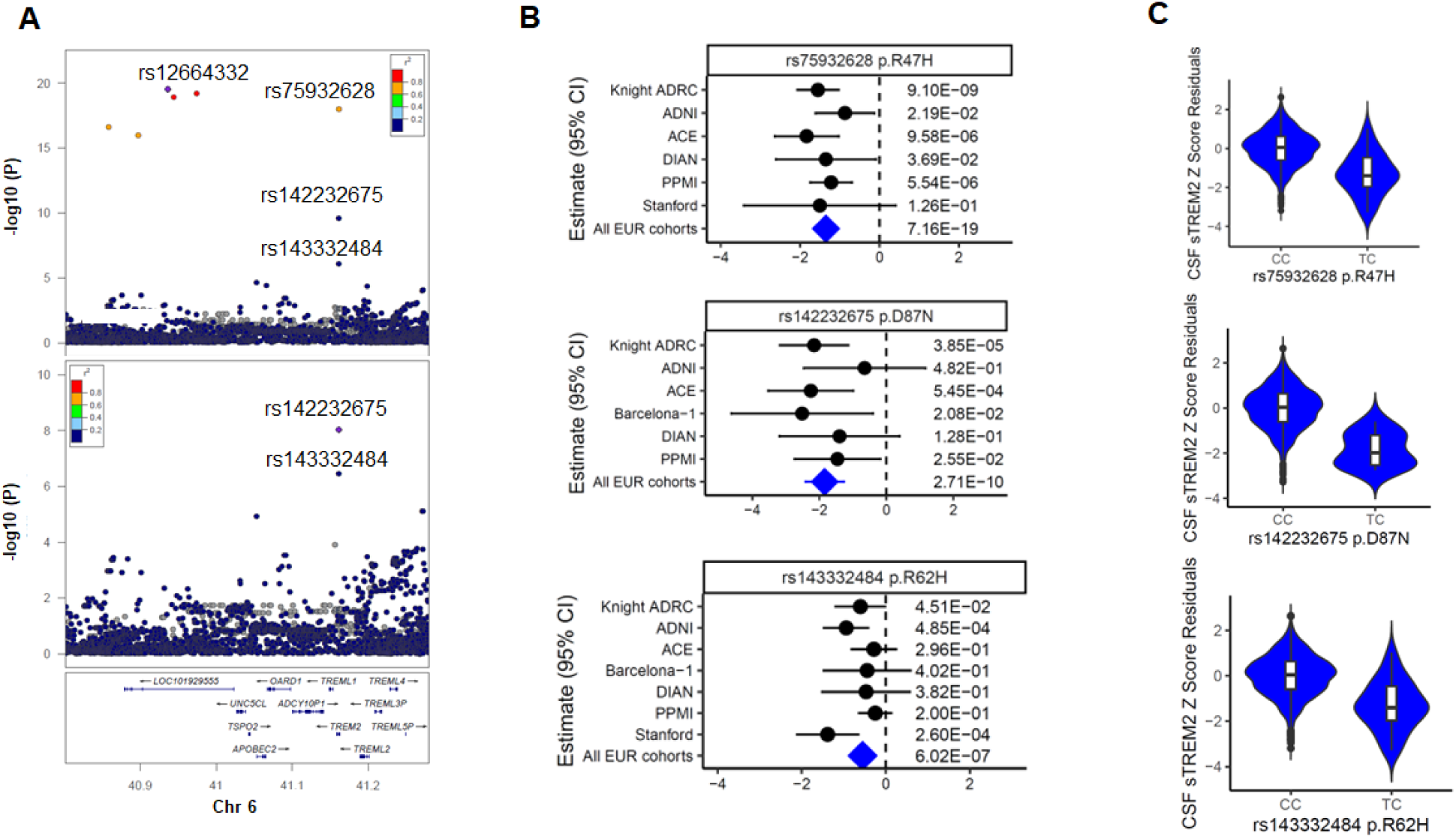
Association results of CSF sTREM2 at chromosome 6. **A)** Top LocusZoom plots at chromosome 6 in European ancestry (EURs) for the sentinel SNP rs12664332 and bottom one is the secondary signal rs142232675 conditioning on the sentinel SNP. X-axis depicts genomic coordinates at chromosome 3 and y-axis denotes the negative log10-transformed P value for each genetic variant. **B)** Forest plots of effect size estimated by cohort for rs142232675 p.D87N, rs75932628 p.R47H, and rs143332484 p.R62H. Heterogeneity P is 5.72 × 10^-1^ for rs75932628, 6.39 × 10^-1^ for rs142232675, and 1.0 × 10^-1^ for rs143332484 respectively. **C)** Violin plots of CSF sTREM2 Z score Residuals vs. genotype of rs142232675 p.D87N, rs75932628 p.R47N, and rs143332484 p.R62H.

### Functional characterization of the chr3p24.1 identifies *TGFBR2* as a novel gene implicated in TREM2 biology

We identified a locus at chromosome chr3p24.1 significantly associated with sTREM2 levels (Fig. 4A and 4B). This locus contained four genetic variants reaching genome-wide significance (Table S3) with consistent homogenous effect size across studies (heterogeneity I^2^ P=0.75; Fig. 4A; Table S3). The sentinel variant (rs73823326, MAF=0.06, β = -0.28, P =3.86×10^-9^) was located between RNA binding motif single stranded interacting protein 3 (*RBMS3*) and transforming growth factor beta receptor 2 (*TGFBR2*).

**Fig. 4.**
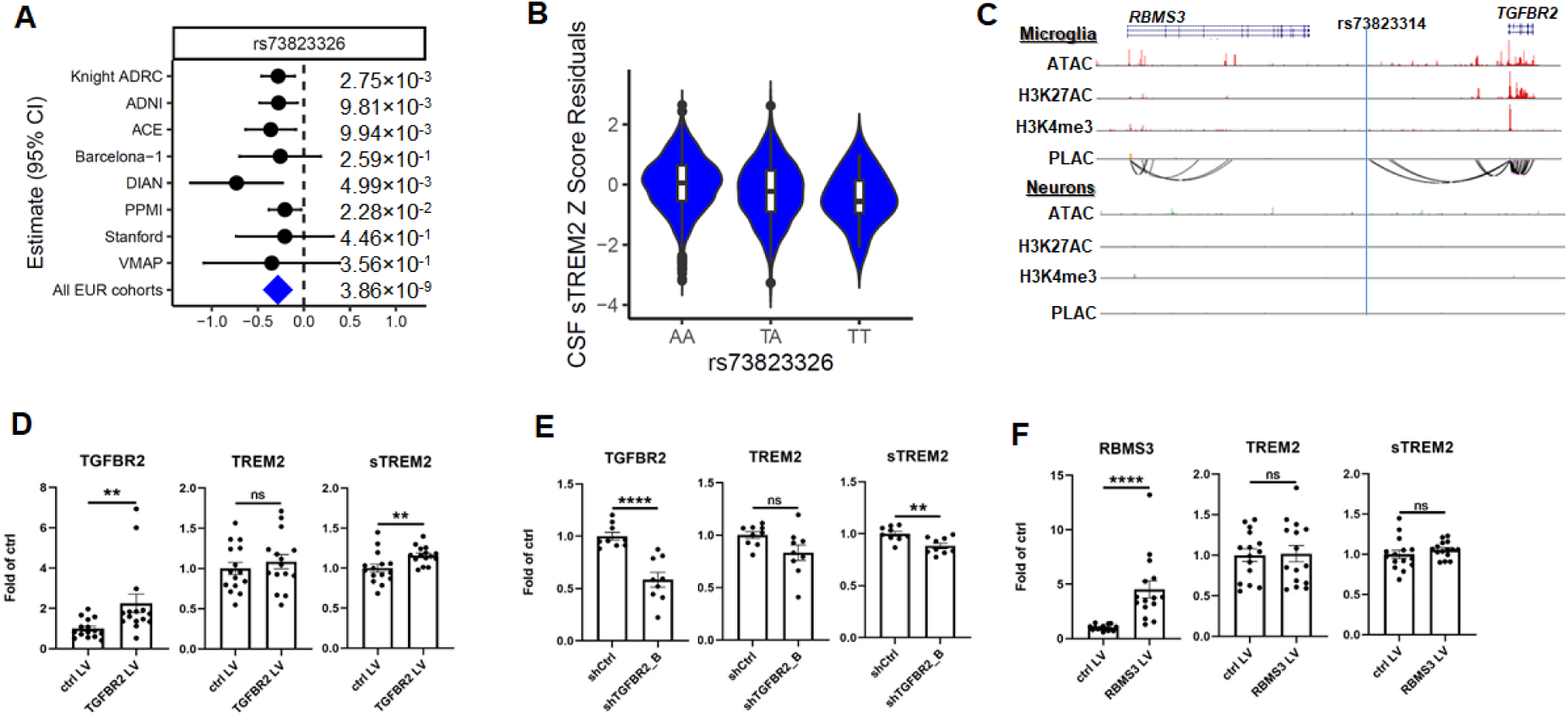
Association results of CSF sTREM2 at chromosome 3 and in vitro functional validation using PBMC-derived macrophages. **A)** Forest plots of effect size estimated by cohort for rs73823326. The effect allele is T for rs73823326. Heterogeneity P is 7.53 × 10^-1^ for rs73823326. **B)** Violin plots of CSF sTREM2 Z Score Residuals by genotypes of rs73823326. **C)** UCSC genome browser visualization of Microglia and Neurons specific assay for transposase-accessible chromatin with sequencing (ATAC-seq), H3K27ac Chromatin immunoprecipitation followed by sequencing (ChiP-seq), H3K4me3 ChiP-seq and proximity ligation-assisted ChIP-Seq (PLAC-seq) loops at the chr 3 *RBMS3 – TGFBR2* locus. Chromatin loops linking the promoter of *TGFBR2* to active gene-regulatory region close to rs73823314 (LD r2=1 with rs73823326 and P=2.54 x 10^-8^) is specific in microglia. **D)** Quantification of intracellular TGFBR2 (left panel), TREM2 (middle panel) and extracellular sTREM2 protein levels in PBMC-derived macrophages upon TGFBR2 overexpression. **E)** Quantification of intracellular RBMS3 (left panel), TREM2 (middle panel) and extracellular sTREM2 protein levels in PBMC-derived macrophages upon RBMS3 overexpression. n = 15 from 4 independent experiments. **F)** Quantification of intracellular TGFBR2 (left panel), TREM2 (middle panel) and extracellular sTREM2 (right panel) protein levels in PBMC-derived macrophages upon TGFBR2 knockdown. n = 9 from 3 independent experiments. ns: not significant, ** p < 0.01, **** p < 0.0001. Results are shown in mean ± SEM.

Among the four genome-wide significant variants, two variants (rs73823314 and rs73823316) were in a regulatory region that binds transcription factor (TF) based on the Ensemble variant effect predictor (VEP) annotation. In order to identify the potential functional variant and gene, we examined brain cell type-specific enhancer-promoter interactome maps [34]. We found high peak of epigenetic markers for *TGFBR2*, consistently measured with ATAC-Seq, H3K27ac, and H3K4me3 (Fig. 4C). These epigenetic markers were only observed in microglia, indicating that *TGFBR2* is actively regulated in microglia. More importantly, we identified microglia-specific dense chromatin loops that connect the regulatory variant rs73823314 (LD with rs73823326, r^2^=1.0) to the promoter of *TGFBR2.* All this evidence suggests that *TGFBR2* is the most likely functional gene that affects CSF sTREM2 levels.

In order to functionally validate these findings, we used human peripheral blood mononuclear cell (PBMC)-derived macrophages as a proxy for microglia (Fig. S7A). To modulate the expression level of these genes within the locus, our first approach was to use lentivirus-mediated overexpression of *TGFBR2* and *RBMS3.* We first confirmed that lentivirus-mediated overexpression increased the protein levels of TGFBR2 (∼2.2-fold, P=0.006) and RBMS3 (∼4.5-fold, P<0.0001) as compared to control lentivirus transduced cells (Fig. 4D and 4E left panels; Fig. S7B). Upon *TGFBR2* overexpression, we observed higher levels of intracellular TREM2 (∼8% increase), although it was not statistically significant (P=0.652; Fig. 4D middle panel, Fig. S7C). This trend was not observed in cells overexpressing *RBMS3* (Fig. 4F middle panel). Importantly, we found a significant increase in extracellular sTREM2 levels upon *TGFBR2* overexpression (∼16% increase, P=0.008; Fig. 4D right panel). We performed the same analysis for *RBMS3* overexpressing cells, but did not observe a change in extracellular sTREM2 levels (Fig. 4F right panel).

To further validate the possible role of *TGFBR2* as a modulator of sTREM2, we used lentivirus-mediated knock-down to reduce the level of *TGFBR2* (Fig. 4E left panel, Fig. S7D and S7E). After confirming a successful silencing of *TGFBR2* by using two independent shRNAs (shTGFBR2_B: ∼42% decrease, P<0.0001; shTGFBR2_D: ∼57% decrease, P=0.001), we saw a trend in lower intracellular TREM2 levels, although it did not reach statistical significance (shTGFBR2_B: ∼16% decrease, P=0.0503; shTGFBR2_D: ∼19% decrease, P=0.093; Fig. 4E middle panel, Fig. S7F). Importantly, silencing of *TGFBR2* led to a significant reduction in extracellular sTREM2 levels (shTGFBR2_B: ∼11% decrease, P=0.0078; shTGFBR2_D: ∼13% decrease, P=0.0078; Fig. 4F right panel, Fig. S7G). Taken together, these data strongly support that *TGFBR2,* not *RBMS3*, is the functional gene in this locus modulating sTREM2 levels.

### The association of the chr19q13.32 genomic region to sTREM2 levels is independent of *APOE*

For the first time, we identified a genome-wide significant association for CSF sTREM2 within the *APOE* (58kb upstream) gene region. There were two variants in LD (r^2^=0.99) reaching genome-wide significance (Fig. 5A) in this region, with consistent association across cohorts (heterogeneity P=0.75; Fig. 5C). The sentinel variant is a common variant (rs11666329, MAF=0.496, β=-0.126, P=2.52×10^-8^) located in an intron of *NECTIN2*. We did not find association of rs11666329 in this locus with CSF APOE2 (P=1.10×10^-1^), CSF APOE3 (P=4.80×10^-1^), or CSF APOE4 (P=8.70×10^-1^).

**Fig. 5.**
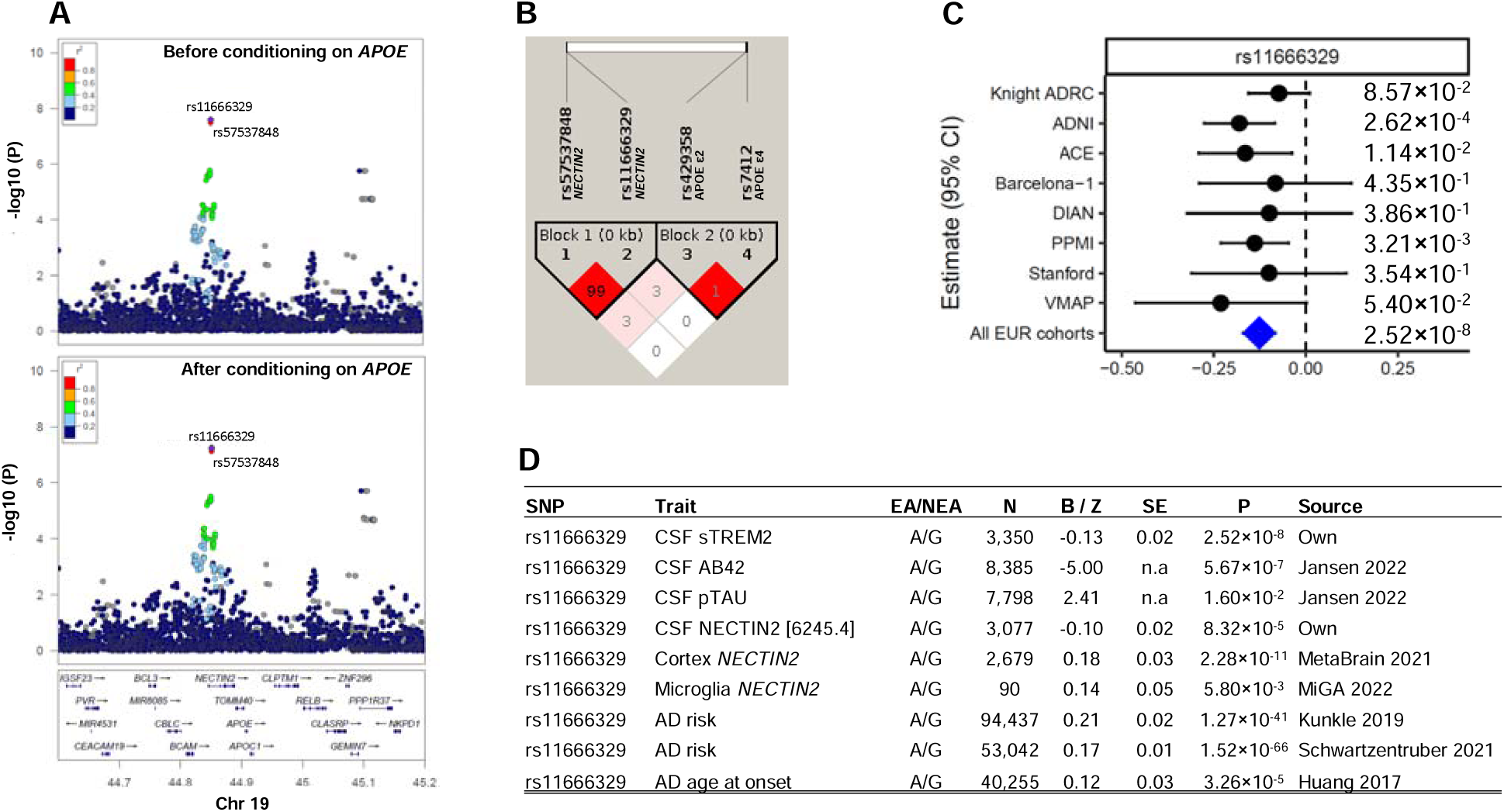
Association results of CSF sTREM2 at chromosome 19. **A)** LocusZoom plots at chromosome 19 before conditional analysis and conditional analyses on Apolipoprotein E (*ApoE*) haplotype. Linkage disequilibrium (LD) estimates used our data. X-axis depicts genomic coordinates at chromosome 19 and y-axis denotes the negative log10-transformed P value for each genetic variant. **B)** Linkage disequilibrium (LD) heatmap of chromosome 19 SNPs and 2 *APOE* snps. **C)** Forest plots of effect size estimated by cohort for rs111666329. The effect allele is G for rs11666329. Heterogeneity P is 7.51 × 10^-1^ for rs11666329. **D)** Association results of rs11666329 for CSF sTREM2, CSF AB42, CSF pTAU, CSF NECTIN2, cortex *NECTIN2*, Microglia *NECTIN2*, AD age at onset, and AD risk.

As *APOE* ε*2* (rs7412) and *APOE* ε*4* (rs429358) are the most significantly associated genetic variants for sporadic AD, we wanted to determine if the association with CSF sTREM2 levels is driven by these known *APOE* variants. We performed conditional analyses for the full *APOE* genotype. However, when we conditioned for the full APOE genotype (or APOE 4 or APOE 2 alone), the association of rs11666329 with CSF TREM2 levels did not change significantly, remaining near genome-wide significant with the similar effect (β=-0.12, P=5.72×10^-8^; Fig. 5A), indicating that this association is independent of *APOE* genotype (Fig. 5A). The LD structure also confirmed that rs11666329 is not in LD with *APOE2* (LD r^2^=0.0031) or *APOE4* (LD r^2^=0.0007; Fig. 5B).

Next, we performed eQTL mapping to determine if *APOE* or any other gene in the region is the most likely functional gene driving this association (Table S10, Table S11 and Table S12). We examined RNA expression of *NECTIN2* (also known as *PVRL2*), *APOE*, *APOC1*, *TOMM40*, and *CLPTM1* gene, across multiple tissues using eQTLGen [35], GTEx [36], Metabrain [37], and microglia (MiGA) [38]. We found that rs11666329 was associated with *NECTIN2* expression in cortex (P=2.78×10^-5^; Metabrain n=2,966; Fig. 5D), with strong evidence of colocalization (PP.H4=0.76; Table S12), as well as with CSF NECTIN2 protein (Fig. 5D). The association of rs11666329 with *NECTIN2* expression was also observed in microglia (P=5.80×10^-3^; MiGA n=90). No such evidence was observed for the other genes in this region (*APOE*, *APOC1*, *TOMM40*, and *CLPTM1*). Brain cell type specific annotation did not observe any interactions between this locus and the promoter of *APOE* (Fig. S8).

We wanted to determine if this variant, rs11666329, is also associated with AD risk, based on the latest GWAS [7, 30]. The A allele of rs11666329 was associated with higher AD risk (β=0.168, P=1.52×10^-66^; Fig. 5D) [7, 30]. In order to address whether the association with AD risk for this variant independent of *APOE* ε2 (rs7412) and ε4 (rs429358), we performed conditional analyses using GCTA-COJO that adjusts for rs7412 and rs429358 in the latest AD GWAS [30]. We observed that the association of rs11666329 with AD risk is still highly significant after conditioning on *APOE* (before conditioning: P=1.52×10^-66^; after conditioning: P=7.12×10^-32^; Table S13), indicating this association is independent of *APOE.* Using the same analyses, the association of rs11666329 with *NECTIN2* expression in cortex is also independent of *APOE* (before conditioning: P=2.78×10^-5^; after conditioning: P=3.32×10^-5^; Table S13).

### Overlap in the genetic architecture of AD risk and CSF sTREM2 levels

In order to determine if the overall genetic architecture of CSF sTREM2 levels overlaps with that of AD risk, beyond the GWAS hits, we determined if polygenic risk scores (PRS) for AD risk (with and without the APOE region) are associated with sTREM2 levels. PRS were computed using effects at genetic variants with P<5.00 ×10^-8^ for AD risk [8]. When variants in *APOE* region were included in PRS calculation, a significant negative association of PRS with CSF sTREM2 was observed (β=-0.047, P=3.57×10^-3^), indicating higher AD PRS has lower CSF sTREM2 level. Since higher AD PRS is with higher AD risk, this negative association further supports the protective effect of CSF sTREM2 level on reducing AD risk. When variants in *APOE* region were removed in PRS calculation, PRS was even more significantly associated with CSF sTREM2 (β=-0.088, P=1.57×10^-7^). Taken together, PRS with AD risk variants has provided significant predictive power for CSF sTREM2.

### Mendelian randomization confirmed the protective role of CSF sTREM2 for AD

To examine whether CSF sTREM2 levels are causal for developing AD, we performed two-sample Mendelian randomization (MR) analysis. We used our GWAS results for CSF sTREM2 and the latest AD GWAS for AD [7]. Eight variants were selected as independent instrument variables after clumping. The variant rs11666329 on chromosome 19 was an outlier noted by MR-PRESSO and removed from this analysis. We chose the remaining seven genetic variants for independent instrument variables (Table S14) and performed five different MR analyses. All analyses provided significant associations, indicating that higher CSF sTREM2 levels lower AD risk (Fig. 6A). The result in MR Egger, which accounts for possible horizontal pleiotropy, remained significant (P=1.78 ×10^-2^). Therefore, we considered the MR results using the inverse variance weighted (IVW) approach as appropriate. Based on this, we conclude that CSF sTREM2 is causal for AD, indicating that higher CSF sTREM2 levels have a significantly protective effect on reducing AD risk (β=-0.236, P=1.36×10^-9^; Fig. 6A and 6B).

**Fig. 6.**
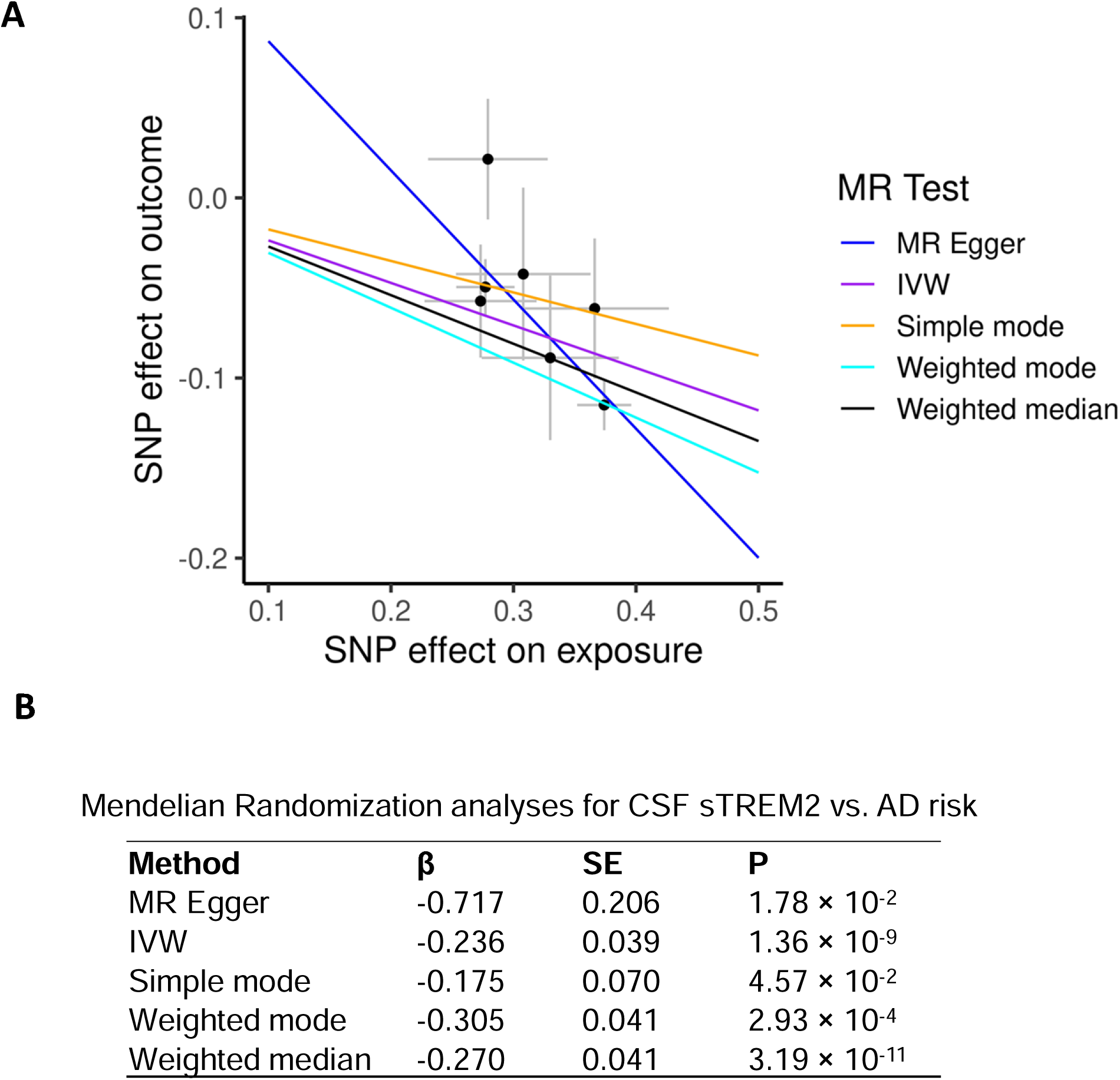
Mendelian randomization (MR) analyses for CSF sTREM2 on AD risk. **A)** Scatter plot of SNP effects on outcome against SNP effects on exposure. The lines represent the causal estimate using 5 method: MR Egger, Inverse variance weighted (IVW), Simple mode, Weighted mode, and Weighted median. **B)** Results of MR analyses for the different MR models

### Understanding the biology of the CSF sTREM2 loci

In order to further characterize the sTREM2 loci, we performed protein-wide association analyses of 7,027 aptamers at the four sentinel variants to identify other proteins that are regulated by the identified loci, using a similar approach that we have used before [39]. We identified three proteins (TREM2, IZUMO4 and A4GALT) associated with rs72918674 on the *MS4A4A/MSA4A6A* locu and 47 (including TREM2, ZNF483, ARL2, and ATE1) with rs11666329 on the *NECTIN2/APOE* locus (Table S15, Fig. S9A, S9B).

IZUMO4 (IZUMO family member 4) is highly expressed in testis and in brain, and genetic variant within its coding gene are associated with height [40], monocyte count [41], neutrophil count [41] and white blood cell count [42]. A4GALT (alpha 1,4-galactosyltransferase) is implicated on the P blood group system. Genetic variants of *A4GALT* are associated with platelet distribution [43], prostate cancer [44], red blood cell count [45], monocyte count [45], and hemoglobin concentration [45]. However at this moment is not clear how these two proteins interact with MS4A4A/MS4A6A or TREM2, and further analyses will be needed. For the 47 proteins associated with the variants located on the *NECTIN2/APOE* locus, we performed pathway enrichment analyses and found significant enriched Amyloid-beta clearance (P=3.81×10^-2^), Innate Immune System (P=5.4×10^-3^), Cytokine Signaling in Immune system (P=1.69×10^-2^), Autophagy (P=4.9×10^-3^), Metabolism of lipids (P=3.81×10^-2^), and Metabolism of proteins (P=3.5×10^-2^), among others (Fig. S9C).

## Discussion

Here, we performed the largest genetic screening for CSF sTREM2 levels integrating protein and genetic data from 3,350 European ancestry individuals, as well as 250 non-Europeans. We identified four genetic loci, including the known *MS4A* cluster on chromosome 11 [27] as well as three novel loci, two of them specific to CSF and not reported in large plasma studies.

We previously reported a *MS4A* cluster as a major regulator for CSF sTREM2 [27]. This study provides additional insights about the role of this locus on AD risk and TREM2. Specifically, we were able to demonstrate that there are two independent signals in the *MS4A* locus and that the *MS4A6A* p.A112T and the *MS4A4A* p.M178V variants are the most likely functional variants based on the multi-ethnic fine mapping analyses. We also demonstrated a significant epistatic effect between these two variants in AD risk and sTREM2 levels. There are other variants in LD with these two variants, some of which are coding or reported to be genetic regulators of gene expression levels (Table S3). It is possible that in each LD blocks the effect is driven by more than one variant, complicating the analyses and interpretation of this region. Additional functional studies will be needed to identify the exact functional variants.

Previously we also demonstrated that genetic and pharmacologic regulation of *MS4A4A* also modify TREM2 levels and therefore *MS4A4A* is a potential therapeutic target for AD. Currently there are several clinical trials that aim to modify sTREM2 levels and AD risk by targeting *MS4A.* For instance, Alector Inc. developed AL044, a humanized MS4A function modulating monoclonal antibody for the treatment of AD (https://investors.alector.com/news-releases/news-release-details/alector-initiates-phase-1-clinical-trial-al044-treatment). In preclinical *in vivo* studies, AL044 has induced key microglial signaling pathways for proliferation, survival, lysosomal activity, migration, phagocytosis, and immune response. The data in this study goes beyond our initial findings and indicating that future investigation of targeting both *MS4A4A* and *MS4A6A* may have even a larger effect than just one of those genes.

*TREM2* risk-variant carriers (rs142232675 p.D87N, rs75932628 p.R47H, and rs143332484 p.R62H) are known to have higher AD risk [7, 8]. With the largest cohort of CSF samples, we were able to detect the association of the *TREM2* locus with CSF sTREM2 levels for the first time. We found significantly lower CSF sTREM2 in *TREM2* risk-variant carriers (rs142232675 p.D87N and rs75932628 p.R47H). The variant p.R47H might reduce CSF sTREM2 level through decreased solubility and cleavage [46]. However, this observation is opposite to the higher CSF sTREM2 in p.R47H variant carriers reported by Piccio et al [21], Deming et al [27], and Suárez-Calvet et al [47]. There are three isoforms of sTREM2 (ENST00000373113, ENST00000373122, and ENST00000338469) in CSF. We do not have the isoform information in Somalogic and ELISA, and the discrepancy between our results and Suárez-Calvet’s might be due to differences in sTREM2 isoforms detected by Somalogic from this study and ELISA from Suárez-Calvet et al [47]. We also cannot rule out the possible influence of p.R47H to sTREM2 structure and sTREM2 binding with the aptamer in the Somalogic platform. Therefore, a follow-up study that quantifies the three isoforms in AD patients and healthy controls using orthogonal proteomic measures including Somalogic, Olink, ELISA, and mass spectrometry would be informative for addressing these possibly conflicting findings.

The novel CSF specific chromosome 3 locus (rs73823326) is located in an intergenic region between *RBMS3* and *TGFBR2*. *TGFBR2* at 3p24.1 encodes a transmembrane protein and plays a key role in signal transduction. Besides hepatic stellate cells, adipocytes and endothelial cells, TGFBR2 is abundant in microglia based on the Human Protein Atlas (https://www.proteinatlas.org). It is reported that brain extracts of AD patients have significantly lower levels of TGFBR2 compared to controls [48]. Consistent with this, reduced neuronal TGFBR2 signaling led to accelerated age-dependent neurodegeneration and promoted beta-amyloid accumulation in an animal model [48]. *TGFBR2* contains a microglia-specific high level of epigenetic markers. The microglia activation induced by Tgfbr2-deficiency [49] further supports a regulatory role of *TGFBR2* in microglia. The presence of the microglia-specific dense chromatin loops that connect this locus to the promoter of *TGFBR2* support the hypothesis that *TGFBR2* is the most likely functional gene underpinning this locus. Since TGFBR2 spans the cell membrane, it might regulate CSF sTREM2 through influencing the proteolytic cleavage at the cell membrane of microglia. Importantly, our *in vitro* cell-based studies confirmed that overexpression (and knock-down) of *TGFBR2* in human primary macrophages increases (and decreases) extracellular sTREM2 levels. We did not observe any changes in sTREM2 levels upon RBMS3 overexpression. Taken together, the bi-directional change on sTREM2 levels due to altered *TGFBR2* expression levels in PBMC-derived macrophages strongly implicates *TGFBR2* as the functional gene in this locus.

Another novel CSF specific signal is a locus (rs11666329) on chromosome 19 located in an intron of *NECTIN2*. Two well-known *APOE* variants (rs429358 and rs7412) reside 50 kb downstream but were not associated with CSF sTREM2. Both LD structure and our conditional analyses confirmed that the association at this locus is independent of *APOE* genotype. Besides CSF sTREM2, the A allele of rs11666329 is associated with significantly lower CSF NECTIN2 protein levels, higher AD risk and earlier age at onset for AD. Active epigenetic markers including ATAC-seq, H3K27ac, and H3K4me3 in this locus were noted in microglia, neurons, and oligodendrocytes. *NECTIN2* is also known as CD112 or PVR-related 2 (PVRL2/PRR2) and its two splice variants (α and δ) are expressed in multiple tissues including brain neurons and astrocytes [50]. Genetic global ablation of *NECTIN2* causes loss of neurons and nerve fibers in mouse brains at 6-months of age [51], indicating a protective role of NECTIN2 in neurodegeneration. Furthermore, one of the SNPs in the human *NECTIN2* gene, is significantly associated with AD in African Americans, even after adjusting for the effects of *APOE* genotype [52]. This is in line with our findings that rs11666329 remains significantly associated with AD after conditioning on two *APOE* variants. Besides sTREM2, additional 46 CSF proteins involved in Amyloid-beta clearance, Innate Immune System, Autophagy, as well as others, were regulated by this locus. Taken together, our analysis is consistent with NECTIN2 being a novel modulator for CSF sTREM2 and that it may impact directly or indirectly AD development.

Our analyses nominate *NECTIN2* as the functional gene for sTREM2 in the *APOE* region but we cannot totally exclude that *APOE* is the functional gene or has no interaction with TREM2 at protein level. APOE has been identified as a binding ligand for TREM2 [53, 54]. The binding of APOE to TREM2 was associated with increased clearance of apoptotic neurons by microglia. Therefore, altered TREM2 protein structure by *TREM2* missense variants, as well as reduced sTREM2 protein levels determined by variants in four loci, might reduce the affinity of APOE for TREM2 and decrease the clearance of beta-amyloid from the brain [53]. This might be one of the possible mechanisms for these loci that contribute to AD risk.

In line with previous results that the soluble form of TREM2 is protective against AD [27]. Our Mendelian randomization and polygenic risks score analyses not only support this hypothesis, but also suggest that the mechanism that regulate the levels of sTREM2 levels is in fact a part of the causal pathway of AD, independently of the *TREM2* risk variants. Our analyses indicate that the variants that regulates sTREM2 levels are also not only regulated AD risk but also slowed memory decline and brain atrophy and reduced amyloid and tau aggregation observed in AD patients [26, 55]. Therefore, modulators of sTREM2 will be ideal candidate for novel AD therapeutic target.

Despite the strength of our study and novel loci identified, the present study has a limitation. A sample size of CSF sTREM2 levels in non-EURs individuals is small. While we narrowed down to the two missense variants for the *MS4A* locus, we were not able to examine the remaining 3 loci. There was not enough power for both chromosome 3 *TGFBR2* locus and chromosome 19 *NECTIN2* locus. In addition, the rare variants in the chromosome 6 *TREM2* locus in Europeans were not available in our non-European cohorts. Follow-up GWAS analyses using non-European population with a larger sample size would be valuable.

In summary, we performed the largest GWAS analysis of CSF sTREM2 and identified four loci. In addition to the known *MS4A* gene cluster and a cis signal in the *TREM2* locus, we identified two novel regulators, *TGFBR2* and *NECTIN2*, involved in TREM2 biology. These two genes may act at the microglia membrane through influencing the proteolytic cleavage and can serve as novel therapeutic targets for AD.

## Material and methods

### Study Design

We performed two-stage GWAS analyses: the first stage of GWAS in joint European ancestry (EURs), and the second stage of multi-ethnic fine mapping. First stage GWAS analyses utilized 3,350 European samples from eight cohorts and 12,621,222 autosomal genotypic variants, and second stage multi-ethnic fine mapping used 250 non-European (non-EURs) samples from eight cohorts and 8,909,120 autosomal genotypic variants. CSF sTREM2 was measured by SomaScan or MSD (Table 1). In the first-stage GWAS analyses, we used an additive linear model adjusting for age at CSF draw, sex, genotype platform/cohorts, and 10 PCs.

For the significant loci, we then conducted post-GWAS analyses. First, we used multi-ethnic fine mapping to detect the true causal variants underlying each locus. For each of the four loci, we then performed stepwise conditional analyses to identify the independent genotypic variants. To identify the functional genes underlying three novel loci, we performed colocalization analyses of each locus with the AD GWAS, GTEx eQTL, and MetaBrain eQTL. The regulatory role of these loci were annotated with the brain cell type-specific enhancer-promoter interaction map. For chromosome 3 RBMS3-TGFBR2 locus, in vitro functional validation using overexpression of TGFBR2 and RBMS3 in human primary macrophages was conducted. The overall genetic architecture overlapped between CSF sTREM2 and AD risk was estimated using association between PRS of AD risk and CSF sTREM2. Finally to determine whether CSF sTREM2 is causal for AD, two-sample Mendelian randomization was analyzed using CSF sTREM2 GWAS as exposure and the latest AD GWAS as outcome.

### Ethics Statement

The Institutional Review Board of all participating institutions approved the study and research was performed in accordance with the approved protocols. Written informed consent was obtained from all participants or their family members.

### Cohort demographics

The join analyses in EURs and non-EURs included participants from Charles F. and Joanne Knight Alzheimer Disease Research Center (Knight ADRC), Alzheimer’s Disease Neuroimaging Initiative (ADNI), ACE Alzheimer Center Barcelona (ACE), Longitudinal observational study from the Memory and Disorder unit at the University Hospital Mutua de Terrassa (Barcelona-1), Dominantly Inherited Alzheimer Network (DIAN), Parkinson’s Progression Markers Initiative (PPMI), Vanderbilt Memory and Aging Project (VMAP) and Stanford ADRC.

Samples were recruited from eight multi-ethnic cohorts. European participants were identified based on principal component analyses (PCA) were used in first stage. In total, 797 samples from Knight ADRC, 676 samples from ADNI, 435 samples from ACE, 187 samples from Barcelona-1, 172 samples from DIAN, and 779 samples from PPMI, 135 participants from VMAP, and 169 individuals from Stanford ADRC were included (Table 1). In the multi-ethnic fine mapping analyses, PCA identified non-European samples including 90 from knight ADRC, 40 from ADNI, 8 from ACE, 6 from Barcelona-1, 31 from DIAN, 38 from PPMI, 7 from VMAP and 30 from Stanford ADRC were analyzed (Table S4).

#### Knight ADRC

Charles F. and Joanne Knight Alzheimer Disease Research Center (Knight ADRC), housed at Washington University in St. Louis, is one of 30 ADRCs funded by NIH. The goal of this collaborative research effort is to advance AD research with the ultimate goal of treatment or prevention of AD. The subjects included in this study are from the Memory and Aging Project (MAP) supported by Knight ADRC. As part of the project, subjects undergo annual psychometric testing and interviews along with biennial or triennial PET, MRI and CSF collection. Further details on Knight ADRC and MAP can be found at https://knightadrc.wustl.edu/. In our discovery stage analyses, 797 EUR samples including 178 (22.33%) of AD cases and 619 (77.67%) cognitive normal controls (hereinafter refers as controls) were from MAP cohort (Table 1). In multi-ethnic fine mapping, a total of 90 non-EURs samples including 12 (13.33%) of AD cases and 78 (86.67%) controls were from MAP cohort (Table S4).

#### ADNI

ADNI was launched in 2003 as a public-private partnership, led by Principal Investigator Michael W. Weiner, MD. The primary goal of ADNI has been to test whether serial magnetic resonance imaging (MRI), positron emission tomography (PET), other biological markers, and clinical and neuropsychological assessment can be combined to measure the progression of mild cognitive impairment (MCI) and early Alzheimer’s disease (AD). For up-to-date information, see www.adni-info.org. EURs samples (n=676) including 512 (75.74%) AD cases and 164 (24.26%) controls were included in discovery stage, non-EURs samples (n=40) including 26 (65%) of AD cases and 14 (35%) controls from ADNI were used in multi-ethnic fine mapping (Table S4).

#### ACE

The ACE study [56] comprises 4120 AD cases and 3289 control individuals. Cases were recruited from ACE Alzheimer Center Barcelona, Institut Català de Neurociències Aplicades (Catalonia, Spain). Diagnoses were established by a multidisciplinary working group, including neurologists, neuropsychologists, and social workers, according to the Diagnostic and Statistical Manual of Mental Disorders–IV criteria for <u>dementia</u> and to the National Institute on Aging and Alzheimer’s Association’s (NIA-AA) 2011 guidelines for defining AD. Control individuals were recruited from three centers: ACE (Barcelona, Spain), Valme University Hospital (Seville, Spain), and the Spanish National DNA Bank Carlos III (University of Salamanca, Spain) (www.bancoadn.org). EURs samples (n=435) including 238 (54.71%) AD cases and 197 controls (45.29%) from ACE were used in discovery stage. non-EURs (n=8) samples including 4 (50%) of AD cases and 4 (50%) of controls were used in multi-ethnic fine mapping (Table S4).

#### Barcelona-1

Barcelona -1 [57] is a longitudinal observational study consisting of ∼300 subjects at baseline carried out in the Memory and Disorder unit at the University Hospital Mutua de Terrassa, Terrassa, Barcelona, Spain. Cases include subjects diagnosed with AD dementia (ADD), non-AD dementias (non-ADD), mild cognitive impairment (MCI), or subjective memory complaints (SMC). Clinical information was collected at baseline as well as longitudinally and lumbar puncture (LP) and amyloid PET were performed if subjects had diagnosis of MCI, early-onset dementia (<65 years), or dementia with atypical clinical features. Our discovery stage in EURs (n=187) included 59 (31.55%) dementia cases and 128 (68.45%) controls from Barcelona-1.The multi-ethnic fine mapping stage in non-EURs included 6 (100%) controls.

#### DIAN

The Dominantly Inherited Alzheimer Network (DIAN), led by Washington University School of Medicine in St. Louis, is focused on the study of Autosomal Dominant AD (ADAD). It is a family-based long-term observational study with standardized clinical and cognitive testing, brain imaging, and biological fluid collection (blood, cerebrospinal fluid) from subjects with the intent of identifying changes in pre-symptomatic and symptomatic gene carriers who are expected to develop AD. Since the focus of this study is on ADAD, which has an early age of onset compared to sporadic AD, the subjects in this cohort are younger on average compared to other cohorts. The data used in this study is from data freeze 15 (DF15). Additional details on DIAN can be found at https://dian.wustl.edu/. 118 (68.6%) ADAD cases and 54 (31.4%) controls were included in discovery stage. Multi-ethnic fine mapping stage included 22 (70.97%) of ADAD cases and 9 (29.03%) of controls.

#### PPMI

The Parkinson’s Progression Markers Initiative (PPMI) [58] is an observational, international study designed to identify clinical, imaging, genetic, and biospecimen Parkinson’s diease (PD) progression markers. This study is a public-private partnership of academic researchers, The Michael J. Fox Foundation for Parkinson’s Research (MJFF), and pharmaceutical and biotech industry partners. The overall goal of PPMI is to investigate novel methods to establish longitudinal PD cohorts to examine clinical, imaging, genetic, and biospecimen PD progression markers that individually or in combination will rapidly demonstrate interval change in PD patients in comparison to Healthy Controls (HC) or in sub-sets of PD patients defined by baseline assessments, genetic mutations, progression milestones, and/or rate of clinical, imaging, or biospecimen change. 460 (59.05%) PD cases and 319 (40.95%) prodromal cases and controls were used in discovery stage. Multi-ethnic fine mapping stage included 27 (71.05) of PD cases and 11 (28.95%) controls.

#### VMAP

The Vanderbilt Memory and Aging Project (VMAP), established in 2012, is a longitudinal study investigating vascular health and brain aging. At baseline, participants complete a physical and frailty examination, fasting blood draw, neuropsychological assessment, echocardiogram, cardiac MRI and brain MRI. The detailed information can be found at https://www.vumc.org/vmac/vanderbilt-memory-aging-project. In discovery stage, 51 (37.78%) dementia cases and 84 (62.22%) controls were included. Multi-ethnic fine mapping stage included three (42.86%) of dementia cases and four (57.14%) of controls.

#### Stanford ADRC

Stanford Alzheimer’s Disease Research Center (ADRC) (https://med.stanford.edu/adrc.html), one of thirty-one ADRC, aims to translate research advances into improved diagnosis and care for people with Alzheimer’s disease and related disorders. The ultimate goals are to prevent and cure AD. 46 (27.22%) dementia cases and 123 (72.78) controls were used in discovery stage. Multi-ethnic fine mapping included 7 (23.33%) of dementia cases and 23 (76.67%) of controls.

### SomaScan and MSD for CSF sTREM2

CSF samples were collected after an overnight fast, processed, and stored at −80 °C for SomaScan assay. CSF sTREM2 for Knight ADRC, ADNI, ACE, Barcelona-1, and DIAN was measured at once using SomaScan v4.1 7K. Whereas the level of CSF sTREM2 for PPMI and Stanford ADRC was assayed separately using SomaScan v4 5K. In-house Meso Scale Discovery (MSD) assay was used to quantify CSF sTREM2 in VMAP cohort [59]. Accordingly, the normalization, quality controls (QCs), and log10 transformation followed by Z score transformation of the cleaned CSF sTREM2 were conducted in 5 cohorts (Knight ADRC, ADNI, ACE, Barcelona-1, and DIAN), PPMI, Stanford ADRC and VMAP.

SomaScan is a multiplexed, single-stranded DNA aptamer-based platform from SomaLogic (Boulder, CO) [60]. Instead of physical units, the protein level was quantified using relative fluorescent units (RFU). To mitigate nuisance variation introduced by the readout, pipetting errors, and inherent sample variation, SomaLogic performed sample level normalization within a plate including hybridization control normalization, intraplate median signal normalization, and median signal normalization to an external reference. The adaptive normalization by maximum likelihood (ANML) was applied for the median signal normalization to an external reference. Finally inter-plate calibration based on calibrator samples was performed to remove plate bias. The details for these normalization procedures were described by Candia J [61].

For both normalized SomaScan readouts and the MSD measures, we further removed outlier datapoints defined as log10 transformed RFU level fell outside of either end of 1.5-fold of interquantile range (IQR). To put the CSF sTREM2 from different platforms onto the same scale, Z score transformation of the values from both SomaScan and MSD was used in final analyses.

### Genotyping and imputation

Genotypes from the eight cohorts were from different platforms. 1) Five different arrays including the Illumina CoreExome-24 (CoreEx), Global Screening Array-24 (GSA), NeuroX2, OmniExpress-24 (OmniEx), and Human660W-Quad (X660W) were used by Knight ADRC. ADNI utlized OmniEx. ACE were genotyped with the Affymetrix Axiom. Genotypes of Barcelona-1 was measured by GSA and NeuroX2. DIAN was genotyped by CoreEx. Only autosomal genetic variants were included in our analyses. The genotypes of these 5 cohorts were quality controlled (including gender check) using PLINK v1.90b6.26 [62] and imputed by our group. Before imputation, the variants and individuals with call rate of <98%, individuals with sex inconsistencies, as well as variants with Hardy-Weinberg equilibrium (HWE) P<1 x 10^-6^, were excluded. The GRCh38/hg38 coordinates based imputation was performed using TOPMed Imputation Server (August 2021). After imputation, imputed variants with Rsq < 0.3 were removed from the data and the hybrid data was created by replacing remaining imputed genotype with actual genotype if available; 2) We obtained whole-genome sequencing data in VCF format (aligned to build GRCh38/hg38) from PPMI. The variants and individuals with call rate of <98% were removed. 3) Whole-genome sequencing data in binary plink format (aligned to GRCh38/hg38) from Stanford ADRC and variants with call rate of <95% and Hardy-Weinberg equilibrium (HWE) P<5 x 10^-8^ were removed by Stanford site. 4) VMAP samples were genotyped on the Illumina Infinium Expanded Multi-Ethnic Genotyping Array (MEGAX) chip on genome build GRCh37. Prior to imputation, Vanderbilt site removed variants with call rate <95% or minor allele frequency (MAF) < 1%. Samples with call rates <99% or exhibited an inconsistency between reported and genetic sex were removed. Variant positions were lifted over to genome build GRCh38 and Imputation was performed on the TOPMed Imputation Server, using Minimac4 and Eagle for phasing. Imputed genetic data were filtered for imputation quality (Rsq>0.8), biallelic SNPs, and MAF >0.01.

The genotypes of the eight cohorts were merged onto one dataset using PLINK v1.90b6.26. We obtained the pairwise genome-wide estimates of proportion identity-by-descent using PLINK v1.90b6.26 [62]. Unanticipated duplicates and cryptic relatedness (Pihat ≥0.20) were identified and the unrelated samples with the higher number of variants were selected. The variants with minor allele count (MAC) > 10 were included in our analyses. The 10 principal components and genetic ancestry (3,350 European and 250 non-European) were calculated using PLINK1.90b6.26 [62]. APOE ε2, ε3, and ε4 isoforms were detected by genotyping rs7412 and rs429358.

### Statistical methods

In first stage GWAS analyses, single-variant association with CSF sTREM2 was conducted jointly for eight cohorts which contain 3,350 European. Additive linear regression model in PLINK v2.0 [62] including age at CSF draw, sex, genotype platforms/cohorts, and 10 principal components were included as covariates. To determine whether the genetic signals demonstrate the consistent effects for CSF sTREM2 across cohorts, we also performed association analyses of each of 8 cohorts separately using the same additive linear regression model accounting for age at CSF draw, sex and 10 principal components.

To identify the independent variants at each locus, we performed stepwise conditional analyses using PLINK v2.0 [62]. In brief, the top SNP of each locus was included as a covariate in the first round, and if any SNP remains significant (P<5 ×10^-8^) after the first round, it will be added in the covariate list. This will be repeated until no significant SNP identified at the locus.

Because of linkage disequilibrium (LD) block, GWAS loci identified in European ancestry contain both causative SNPs and the variants in LD with them. Leveraging the fact that the population evolution in different ethnic group creates allelic heterogeneity and LD block variations, cross-ancestry fine-mapping can be useful for pin-pointing the likely functional variants. The GWAS signals shared by multi-ancestry are more likely the functional causal variants. To achieve this goal, we performed additive linear regression model for 250 non-European participants jointly. Since the number of non-Europeans in each cohort does not have sufficient power, we did not analyze eight cohorts separately.

Finally, A fixed effect meta-analyses of EURs and non-EURs was performed using METAL [63]. The significant signals were determined as 1) P< 5 × 10^-8^ in stage 1; 2) P<0.05 in stage 2 and the concordant direction of effect estimation as in stage 1; 3) P< 5 × 10^-8^ in trans-ancestry meta-analyses and the concordant direction of effect estimate between stage 1 and stage 2.

Regional visualization of GWAS results were generated using LocusZoom v1.3 [64] and forest plots were produced by ggplot2 in R v3.5.2. Heat map of pairwise linkage disequilibrium (LD) was produced using Haploview [65]. The pairwise interactions among SNPs were performed using R (version 4.2.1).

For chromosome 19 APOE locus, to identify whether our variants are independent of *APOE*, *APOE* ε2, ε3, and ε4 isoforms were determined by genotyping rs7412 and rs429358 using Taqman genotyping technology. We coded the *APOE* haplotype as 0 for ε2/ε2, 1 for ε2/ε3, 2 for ε3/ε3, 3 for ε2/ε4, 4 for ε3/ε4, and 5 for ε4/ε4 and included this as a covariate in PLINK v2.0 [62].

To identify whether our chromosome 19 variants is significant for AD and *NECTIN2* after adjusting for two APOE SNPs, we downloaded the AD GWAS[30] and MetaBrain eQTL [37] and performed conditioning analyses using GCTA [66].

### Bioinformatics annotation

The rsID number, genes affected by our variants, consequence of our variants on the protein sequence, and location of the variants related to genes were annotated with the Ensemble Variant Effect Predictor (VEP) release 107 (GRCh38.p13 assembly for Homo_sapiens) [67]. If the top signals were also expression quantitative trait loci (eQTLs) or protein quantitative trait loci (pQTLs) and associated with particular gene or protein expression, the gene or the protein is most likely the functional molecules underlying the locus. For this purpose, expression quantitative trait loci (eQTL) from Genotype-Tissue Expression (GTEx) Analysis V8 [36], MetaBrain [37], and blood eQTLGen [35] were used to assist in functional annotation. Additional microglia specific eQTL from MiGA [38] were also used to annotate variants. We also utilized our internal pQTLs for proteins assayed in SomaScan 7K to annotate and interpret our findings. The UCSC genome browser session (hg19) containing the processed ATAC-seq, ChIP-seq, and PLAC-seq datasets for each brain cell type was used for the brain cell type specific enhancer-promoter interaction map annotation [34].

### Mendelian Randomization

To investigate the role of CSF sTREM2 in AD risk, we conducted Mendelian randomization (MR) analyses. MR has been widely used to determine the causal relations between modifiable exposures and disease. We used two-sample MR implemented in R package TwoSampleMR [68] version 0.5.5 to test whether CSF sTREM2 is causal for AD risk. The latest AD GWAS results [8] were treated as disease outcome. First, non-palindromic SNPs with association P < 5 ×10^-8^ for CSF sTREM2 were selected and independent genetic variants was identified using clump_kb=10000 and clump_r2=0.1. These variants were used as instruments and were extracted from AD GWAS results (Table S14). After data harmonization step, two-sample MR applied MR Egger, Weighted median, Inverse variance weighted (IVW), Simple mode and Weighted mode to estimate the causal effect of CSF sTREM2 on AD risk. Additionally we performed MR pleiotropy residual sum and outlier (MR-PRESSO) tests. We used the following strategy to select the most appropriate MR tests. 1) If global horizontal pleiotropy was confirmed by MR-PRESSO, the outlier SNPs were removed from the instrument lists and corrected P was selected from MR-PRESSO; 2) If there was no evidence of global horizontal pleiotropy and there was significant egger intercept, MR-Egger test was selected; 3) If both global horizontal pleiotropy and egger intercept were not significant, inverse variants weighted meta-analysis (IVW), aggregating all of single-SNP causal effects, was used instead. P<0.05 in MR analyses was considered significant.

### Co-localization analyses

To investigate whether CSF sTREM2 and AD risk and relevant genes shared the same causal genetic architecture, we performed bayesian factor colocalization analyses using coloc.abf function in coloc R package (version 5.1.1) [69]. AD GWAS results by Bellenguez C et al [8] were based on 111,326 clinically diagnosed/proxy AD cases and 677,663 controls. Cis-eQTL datasets were obtained for whole blood [35] (https://www.eqtlgen.org), 54 GTEx tissues [36] (release v.8, dbGaP: phs000424.v8) and MetaBrain (https://www.metabrain.nl).This package first performed fine-mapping under a single causal variant assumption, then integrating over two posterior distribution to calculate the probabilities that these variants were shared. The default priors as P1=1×10^-4^, P2=1×10^-4^ and P12=1×10^-5^ were used in our analyses. There are 5 posterior probabilities: PP.H0, PP.H1, PP.H2, PP.H3, and PP.H4. H0 indicates neither trait has a genetic association in the region. H1 is for trait 1 has a genetic association in the region. H2 indicates only trait 2 has a genetic association in the region. H3 indicates both traits are associated, but with different causal variants. H4 represents both traits are associated and share a single causal variant. PP.H4 > 0.8 was used to declare that two traits share the same causal variants.

### Polygenic risk score (PRS) analysis

To calculate PRS for AD, we downloaded the summary statistics of the largest GWAS study for AD [8]. The PRSice-2 was then used to calculate the PRS [70]. First, PRSice-2 utilized “C+T” method to choose the AD risk variants that is clumping and retaining independent SNPs with the smallest P in a 250-kb window according to LD r^2^<0.1, as well as P thresholding such as 5×10^-8^, 5×10^-5^, 0.05 and 0.5. The AD risk variants in our analyses have P<5×10^-8^. Second, using effects of these AD risk variants as weights, PRS for AD was calculated as weighted sum of the risk allele for our samples. Finally, multivariate linear regression was used to assess the association of this PRS and CSF sTREM2 accounting for age at CSF draw, sex, genotype platforms/cohorts, and 10 principal components.

### Tissue Specificity of Identified genetic loci

To determine whether the identified genetic loci are CSF specific, we examined the association of these loci with sTREM2 in plasma based on 35,559 Icelanders (Table S2) [71].

### Sensitivity analyses of impacts of disease status

CSF amyloid beta42 (Aβ42) and phosphorylated tau-181 (pTau) levels were used to classified samples into biomarker negative (A-T-) and positive (A+T+) by Mclust function “mclust” R package (version 5.4.6) via Gaussian mixture models fitted by expectation-maximization algorithm. Z-score of the log10-transformed Aβ42 and pTau for each cohort was used to determine the outliers that were outside of 3SD of the mean. After outlier removal, z-score was re-estimated and used for classification separately for five cohorts (shown below). Due to missing CSF Aβ42 and pTau in PPMI, we did not include PPMI in our sensitivity analyses.

In MAP, both CSF Aβ42 and pTau were measured using the LumiPulse G platform (Fujirebio US, Inc, Malvern, PA). Since there is high correlation between Innotest (Fujirebio) and LumiPulse G platform (r=0.73 for AB42 and r=0.86 for pTau), Innotest values were used for 17 samples due to missing LumiPulse measures. A total of 948 subjects were included for classification. A cutoff of z-score = -0.20 was obtained for Aβ42 corresponding to a raw value of 630 pg/mL. Samples below 630 were considered Aβ42 positive. A cutoff of z-score = 0.61 was obtained for pTau corresponding to a raw value of 62.9. Samples above 62.9 were considered pTau positive.

In ADNI, CSF Aβ42 was assayed in Innotest (Fujirebio) and pTAU was measured using Elecsys (F. Hoffmann-La Roche Ltd, Switzerland). A total of 749 subjects were included for dichotomization. A z-score cutoff of 0.616 was identified for Aβ42, corresponding to a raw value of 196 pg/mL. Samples below 196 were considered Aβ42-positive. A z-score cutoff of 0.197 was identified for pTau, corresponding to a raw value of 27.8. Samples above 27.8 were considered to be pTau-positive.

In Barcelona-1, both CSF Aβ42 and pTau were measured using Innotest for 231 samples. A z-score cutoff of 1.04 was identified for Aβ42, corresponding to a raw Aβ42 value of 1325 pg/mL. Samples below 1325 pg/mL were considered Aβ42-positive. A z-score cutoff of -0.163 was identified for pTau, corresponding to a raw value of 58. Samples above 58 were considered to be pTau-positive.

In ACE, both CSF Aβ42 and pTau were measured using Innotest for 632 samples. A z-score cutoff of 0.468 was identified for Aβ42, corresponding to a raw Aβ42 value of 856 pg/mL. Samples below 856 were considered Aβ42-positive. A z-score cutoff of -0.018 was identified for pTau, corresponding to a raw value of 67. Samples with a value greater than 67 were considered pTau-positive.

In DIAN, LumiPulse was used for both Aβ42 and pTau. Due to missingness, Five Aβ42 values and ten pTau values were replaced with CSF_xMAP (MilliporeSigma, Burlington, MA) platform measurements (r=0.77 and 0.89 between platforms for Aβ42 and pTau respectively). Dichotomization for Aβ42 was performed on 478 samples and 474 for pTau. A z-score cutoff of - 0.198 was identified for Aβ42, corresponding to a raw Aβ42 value of 517 pg/mL. Samples below 517 were considered to be Aβ42-positive.

To examine the impact of disease status on our signals, we performed the following sensitivity analyses:

1) Association analyses with age at CSF draw, sex, genotype platforms/cohorts, 10 principal components, and AD status (AD coded as 1 and CO coded as 0; n=1,972) as covariates;
2) Association analyses with age at CSF draw, sex, genotype platforms/cohorts, 10 principal components, and AT classification (A+T+ coded as 1 and A-T-coded as 0) (n=1,600) as covariates;
3) Association analyses with age at CSF draw, sex, genotype platforms/cohorts, 10 principal components, and clinical dementia rating (CDR) (n=1,639) as covariates;
4) Association analyses with age at CSF draw, sex, genotype platforms/cohorts, and 10 principal components as covariates for biomarker negative (A-T-) individuals (n=841);
5) Association analyses with age at CSF draw, sex, genotype platforms/cohorts, and 10 principal components as covariates for biomarker positive (A+T+) individuals (n=759).

### PheWAS analyses of four identified genetic loci

For the four sentinel variants, we performed protein-wide association analyses for 7,027 aptamers using additive linear regression model in PLINK v2.0 [62] with age at CSF draw, sex, genotype platforms/cohorts, and 10 principal components as covariates. The significant associations were defined by P< 5 × 10^-8^. Additional pathway enrichment analyses were conducted using Enrichr (https://maayanlab.cloud/Enrichr/) with the genes assayed in SomaScan 7K as backgrounds.

### Isolation, culture and differentiation of peripheral blood mononuclear cells (PBMCs)

PBMCs were isolated immediately after blood draw via density gradient centrifugation using Ficoll-Paque™ PLUS reagent (17144003, Cytiva/Thermo Scientific, Waltham, MA, USA). Briefly, blood was removed from anticoagulant (ethylenediaminetetraacetic acid (EDTA)) containing collection tubes and diluted 1:1 with sterile phosphate buffered saline (PBS; 14190-136, Gibco). Diluted blood was carefully pipetted into sterile 50 ml conical tubes containing 15 ml of Ficoll reagent and centrifuged at 1200 x g for 30 min at RT. PBMCs at the interphase were carefully removed and transferred into new 50 ml conical tubes and washed twice with PBS by centrifugation at 500 x g for 10 min at 4°C. PBMCs were plated in non-supplemented RPMI 1640 (11875-085, Gibco/ Thermo Scientific) medium and incubated for 2 h in humidified incubator maintained at 37°C and 5% CO_2_. Cells were washed twice with PBS and started the differentiation into macrophages by adding RPMI 1640 medium containing 10% HyClone defined fetal bovine serum (FBS; SH30070.03HI, Cytiva), 2 mM L-Glutamine (25030081, Gibco), 100 U/ml penicillin and 100 μg/ml streptomycin (15140-122, Gibco), 1X MEM nonessential amino acids, 31.25 mM HEPES buffer, 1 mM Sodium pyruvate (25-025-CI, 25-060-CI and 25-000-CI, Corning, Corning, NY, USA), and 50 ng/ml of recombinant human macrophage colony stimulating factor (M-CSF; 300-25, Peprotech/Thermo Scientific) to cells. Full medium change was performed on 3 and 7 days in vitro (DIV). Cells and conditioned medium were harvested on DIV 9.

### Lentiviral transduction

PBMC-derived macrophages were transduced at DIV 3 after medium change by adding the lentivirus into the cell culture medium. Multiplicity of infection 1 (MOI 1) was used for all lentiviruses. The following lentiviruses were used: TGF beta Receptor II (NM_001024847) Human Tagged ORF Clone Lentiviral Particle (RC223209L3V), RBMS3 (NM_001003793) Human Tagged ORF Clone Lentiviral Particle (RC211441L3V), Lentiviral Control Particles (PS100092V), TGF beta Receptor II Human shRNA Lentiviral Particle (shRNAs TL308851VB and TL308851VD; Locus ID 7048, TL308851V), Lentiviral shRNA Control Particles (TR30021V). The cDNAs and shRNAs were in pLenti-C-Myc-DDK-P2A-Puro and pGFP-C-shLenti plasmids, respectively. All the lentiviruses were obtained from OriGene Technologies (Rockville, MD, USA).

### Immunoblotting

Cells were lysed in ice-cold M-PER™ Mammalian Protein Extraction Reagent (78501, Thermo Scientific, Waltham, MA, USA) supplemented with protease and phosphatase inhibitors (A32965 and A32957, Thermo Scientific). The samples were incubated on ice for 20 min and centrifuged at 16,000 x g for 10 min at 4°C. Protein concentration was determined using Pierce BCA Protein Assay Kit (23227, Thermo Scientific) according to manufacturer’s instructions. Samples were prepared using XT sample buffer and XT reducing agent (1610791 and 1610792, Bio Rad, Hercules, CA, USA) and boiled for 5 min at 95°C. Ten to 20 ug of total protein were loaded per lane and separated on SDS-PAGE using 4-12% Criterion XT Bis-Tris gels (3450123 and 3450124, Bio Rad), and transferred onto PVDF membrane (1620177, Bio Rad) using semi-dry Trans-Blot® Turbo™ protein transfer system (1704150, Bio Rad). Next, membranes were blocked for 1 h at RT in 5% skimmed milk in Tris buffered saline with 0.05% Tween-20 (P7949, MilliporeSigma, St. Louis, MO, USA) (TBST). All primary antibody incubations were performed on a shaker overnight at 4°C. The following primary antibodies and dilutions were used: recombinant anti-TGF beta Receptor II antibody [EPR14673] (1:1000, ab184948, Abcam, Waltham, Boston, MA, USA), anti-RBMS3 (1:1000, NBP1-89497, Novus biologicals, Englewood, CO, USA), anti-TREM2 (AF1828, R&D Systems, Minneapolis, MN, USA), anti-GAPDH (1:4000, MA5-15738, Thermo Scientific) and anti β-actin (1:5000, MilliporeSigma, St. Louis, USA). After removing the primary antibodies, the membranes were washed 3 x 5 min with TBST at RT followed by secondary antibody incubation with horseradish peroxidase-conjugated anti-mouse, anti-rabbit IgG (7076S and 7074S, Cell Signaling Technology) or anti-goat IgG (A27014, Thermo Scientific). All secondary antibodies were diluted 1:5000 in 5% skimmed milk in TBST and incubated for an hour at room temperature. Signals were visualized using either Clarity Western ECL Substrate (1705061, Bio Rad) or SuperSignal™ West Femto Maximum Sensitivity Substrate (34095, Thermo Scientific). The blots were imaged using ChemiDoc^TM^ Imaging System (12003153, Bio Rad) and band intensity was quantified using ImageJ software (National Institutes of Health). Signals were normalized to GAPDH or β-actin signal, which were used as loading control. The values shown are the normalized band intensities relative to the experimental control group. Quantifications of western blots were performed by using Fiji software.

### Enzyme-linked immunosorbent assay (ELISA)

Conditioned medium was harvested on DIV 9 from PBMC-derived macrophages. Conditioned medium was centrifuged at 2000 x g for 10 min at 4°C to remove cells and membrane debris. Next, conditioned medium was transferred into fresh tubes and stored at -80°C until used for ELISA. Human TREM2 ELISA kits (EH464RB, Thermo Scientific) were used to measure soluble TREM2 levels in conditioned medium. All the samples were run in duplicates in each assay and each batch was normalized to its respective controls.

### Statistical analyses for cell-based experiments

Statistical analyses were performed using non-parametric unpaired t test (Mann-Whitney test) using GraphPad Prism 8 software (GraphPad Software, La Jolla, CA). Statistical significance threshold of p < 0.05 was used. ns: not significant, * p < 0.05, ** p < 0.01, *** p < 0.001, **** p < 0.0001. Mean ± SEM per group are shown.

## Data Availability

Data from Knight ADRC participants can be accessed at https://www.niagads.org/knight-adrc-collection. Data from DIAN, ADNI and PPMI can be found at https://dian.wustl.edu/our-research/for-investigators/diantu-investigator-resources/dian-tu-biospecimen-request-form/, https://adni.loni.usc.edu/, and https://www.ppmi-info.org/ respectively.

## Abbreviations

TREM2: Triggering receptor expressed on myeloid cells 2
sTREM2: Soluble TREM2
CSF: Cerebrospinal fluid
GWAS: genome-wide association study
AD: Alzheimer’s disease
EURs: European ancestry
Non-EURs: non-European individuals
LD: Linkage disequilibrium
TF: Transcription factor
VEP: Ensemble variant effect predictor
PBMC: Peripheral blood mononuclear cell
eQTL: expression quantitative trait loci
MR: Mendelian randomization
Knight ADRC: Charles F. and Joanne Knight Alzheimer Disease Research Center
ADNI: Alzheimer’s Disease Neuroimaging Initiative (ADNI)
ACE: ACE Alzheimer Center Barcelona
Barcelona-1: Longitudinal observational study from the Memory and Disorder unit at the University Hospital Mutua de Terrassa
DIAN: Dominantly Inherited Alzheimer Network
PPMI: Parkinson’s Progression Markers Initiative
VMAP: Vanderbilt Memory and Aging Project

## Acknowledgements

This work was supported by access to equipment made possible by the Hope Center for Neurological Disorders, the NeuroGenomics and Informatics Center (NGI: https://neurogenomics.wustl.edu/) and the Departments of Neurology and Psychiatry at Washington University School of Medicine.

We thank all the participants and their families, as well as the many involved institutions and their staff.

This work was supported by access to equipment made possible by the Hope Center for Neurological Disorders, the Neurogenomics and Informatics Center (NGI: https://neurogenomics.wustl.edu/) and the Departments of Neurology and Psychiatry at Washington University School of Medicine.

ADNI resources: Data used in preparation of this article were obtained from the Alzheimer’s Disease Neuroimaging Initiative (ADNI) database (adni.loni.usc.edu). As such, the investigators within the ADNI contributed to the design and implementation of ADNI and/or provided data but did not participate in analysis or writing of this report. A complete listing of ADNI investigators can be found at: http://adni.loni.usc.edu/wp-content/uploads/how_to_apply/ADNI_Acknowledgement_List.pdf. Data collection and sharing for this project was funded by the Alzheimer’s Disease Neuroimaging Initiative (ADNI) (National Institutes of Health Grant U01 AG024904) and DOD ADNI (Department of Defense award number W81XWH-12-2-0012). ADNI is funded by the National Institute on Aging, the National Institute of Biomedical Imaging and Bioengineering, and through generous contributions from the following: AbbVie, Alzheimer’s Association; Alzheimer’s Drug Discovery Foundation; Araclon Biotech; BioClinica, Inc.; Biogen; Bristol-Myers Squibb Company; CereSpir, Inc.; Cogstate; Eisai Inc.; Elan Pharmaceuticals, Inc.; Eli Lilly and Company; EuroImmun; F. Hoffmann-La Roche Ltd and its affiliated company Genentech, Inc.; Fujirebio; GE Healthcare; IXICO Ltd.; Janssen Alzheimer Immunotherapy Research & Development, LLC.; Johnson & Johnson Pharmaceutical Research & Development LLC.; Lumosity; Lundbeck; Merck & Co., Inc.; Meso Scale Diagnostics, LLC.; NeuroRx Research; Neurotrack Technologies; Novartis Pharmaceuticals Corporation; Pfizer Inc.; Piramal Imaging; Servier; Takeda Pharmaceutica l Company; and Transition Therapeutics. The Canadian Institutes of Health Research is providing funds to support ADNI clinical sites in Canada. Private sector contributions are facilitated by the Foundation for the National Institutes of Health (www.fnih.org). The grantee organization is the Northern California Institute for Research and Education, and the study is coordinated by the Alzheimer’s Therapeutic Research Institute at the University of Southern California. ADNI data are disseminated by the Laboratory for Neuro Imaging at the University of Southern California.

*DIAN resources:* Data collection and sharing for this project was supported by The Dominantly Inherited Alzheimer Network (DIAN, U19AG032438) funded by the National Institute on Aging (NIA),the Alzheimer’s Association (SG-20-690363-DIAN), the German Center for Neurodegenerative Diseases (DZNE), Raul Carrea Institute for Neurological Research (FLENI), Partial support by the Research and Development Grants for Dementia from Japan Agency for Medical Research and Development, AMED, and the Korea Health Technology R&D Project through the Korea Health Industry Development Institute (KHIDI), Spanish Institute of Health Carlos III (ISCIII), Canadian Institutes of Health Research (CIHR), Canadian Consortium of Neurodegeneration and Aging, Brain Canada Foundation, and Fonds de Recherche du Québec – Santé. This manuscript has been reviewed by DIAN Study investigators for scientific content and consistency of data interpretation with previous DIAN Study publications. We acknowledge the altruism of the participants and their families and contributions of the DIAN research and support staff at each of the participating sites for their contributions to this study.

ACE Alzheimer Center Barcelona acknowledges all patients and their families for their collaboration. For CSF biomarker research, LA.R. and M.B. received support from the European Union/EFPIA Innovative Medicines Initiative Joint undertaking ADAPTED and MOPEAD projects (grant numbers 115975 and 115985, respectively). M.B. and A.R. are also supported by national grants PI13/02434, PI16/01861, PI17/01474, PI19/01240,PI19/01301, PI22/01403 Lfrom the Acción Estratégica en Salud, integrated in the Spanish National RCDCI Plan and funded by Instituto de Salud Carlos III (ISCIII)—Subdirección General de Evaluación and the Fondo Europeo de Desarrollo Regional (FEDER—“Una manera de Hacer Europa”). A.R. and M.B. have also received support from CIBERNED (Instituto de Salud Carlos III (ISCIII). A.R. is also supported by the EXIT project, EU Euronanomed3 Program JCT2017, Grant No. AC17/00100 and PREADAPT project; the Joint Program for Neurodegenerative Diseases (JPND), Grant No.LAC19/00097; Acción Estratégica en Salud, integrated in the Spanish National RCDCI Plan and funded by Instituto de Salud Carlos III (ISCIII)—Subdirección General de Evaluación and the Fondo Europeo de Desarrollo Regional (FEDER—“Una manera de Hacer Europa”).LI. de Rojas is supported by a national grant from the Instituto de Salud Carlos III FI20/00215.

VMAP data was obtained from the Vanderbilt Memory and Aging Project. Data were collected and processed by Vanderbilt Memory and Alzheimer’s Center investigators at Vanderbilt University Medical Center funded by the following grants: R01-AG034962, R01-AG056534, R01-NS100980, R01-AG062826, and Alzheimer’s Association IIRG-08-88733. Additional funding for cerebrospinal fluid processing from the Swedish Research Council #2018-02532, the European Research Council #681712, and the Swedish State Support for Clinical Research #ALFGBG-720931. Genotyping supported by R01-AG059716.

## Funding

This work was supported by grants from the National Institutes of Health (R01AG044546 (CC), P01AG003991 (CC, JCM), RF1AG053303 (CC), RF1AG058501 (CC), U01AG058922 (CC), RF1AG074007 (YJS)), the Chan Zuckerberg Initiative (CZI), the Michael J. Fox Foundation (CC), the Department of Defense (LI-W81XWH2010849), and the Alzheimer’s Association Zenith Fellows Award (ZEN-22-848604, awarded to CC).

The recruitment and clinical characterization of research participants at Washington University were supported by NIH P30AG066444 (JCM), P01AG03991 (JCM), and P01AG026276 (JCM).

## Author contributions

Conceptualization: CC, YJS; Methodology: LW, NPN, DW, YJS, and CC involved in the analyses; PG and JT involved in data quality control; Original draft: LW, NPN, YJS, CC; Review & editing: All authors reviewed, edited and approved the manuscript.

## Competing interests

CC has received research support from: GSK and EISAI. The funders of the study had no role in the collection, analysis, or interpretation of data; in the writing of the report; or in the decision to submit the paper for publication. CC is a member of the advisory board of Vivid Genomics and Circular Genomics and owns Stocks. TH is on the Scientific Advisory Board for Vivid Genomics. AG is a member of the Scientific Advisory Board for Genentech and Muna Therapeutics.

## Consent to publication

All authors have read and approved the manuscript submitted.

## Ethics Declarations

All participants provided informed consent for their data to be used in this study. The study was approved by the institutional review board of Washington University School of Medicine in St.Louis.

## Supplementary information

The PDF file includes:

Fig. S1 to S9

## Other Supplementary Materials for this manuscript included the following

Tables S1 to S15

## Supplementary figures

**Fig. S1.**
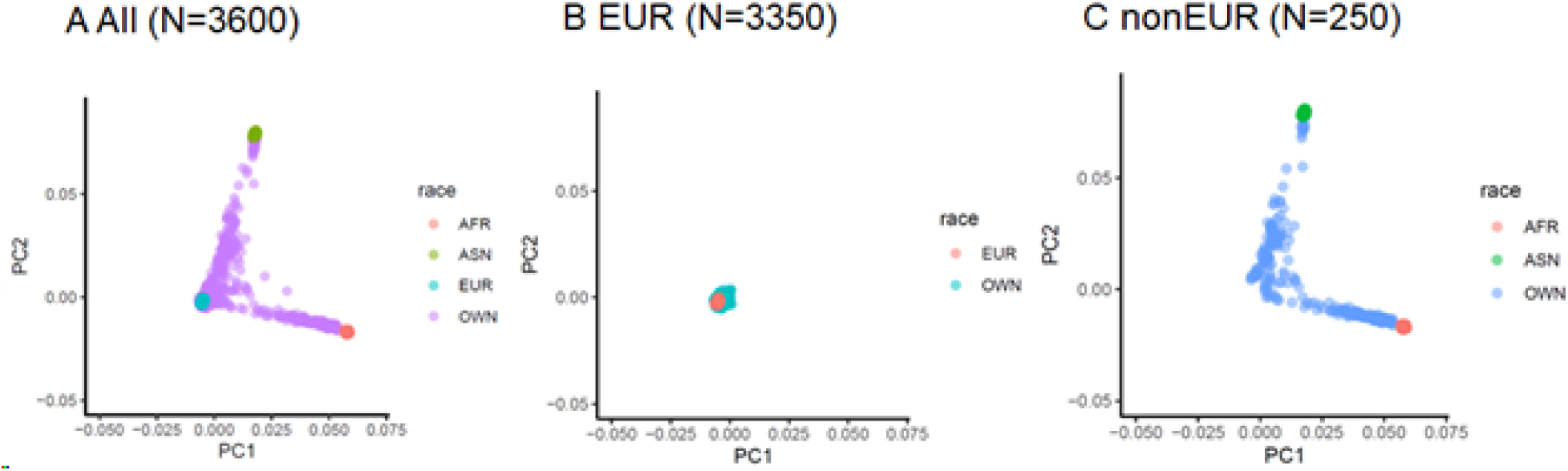
Principal component plots of C2 vs C1 using HapMap Phase II as reference. A) PCA plots for all samples (N=3600). B) PCA plots for EURs (N=3350). C) PCA plots for nonEURs (N=250). AFR indicates Africans; ASN denotes Asian; EUR indicates Europeans.

**Fig. S2.**
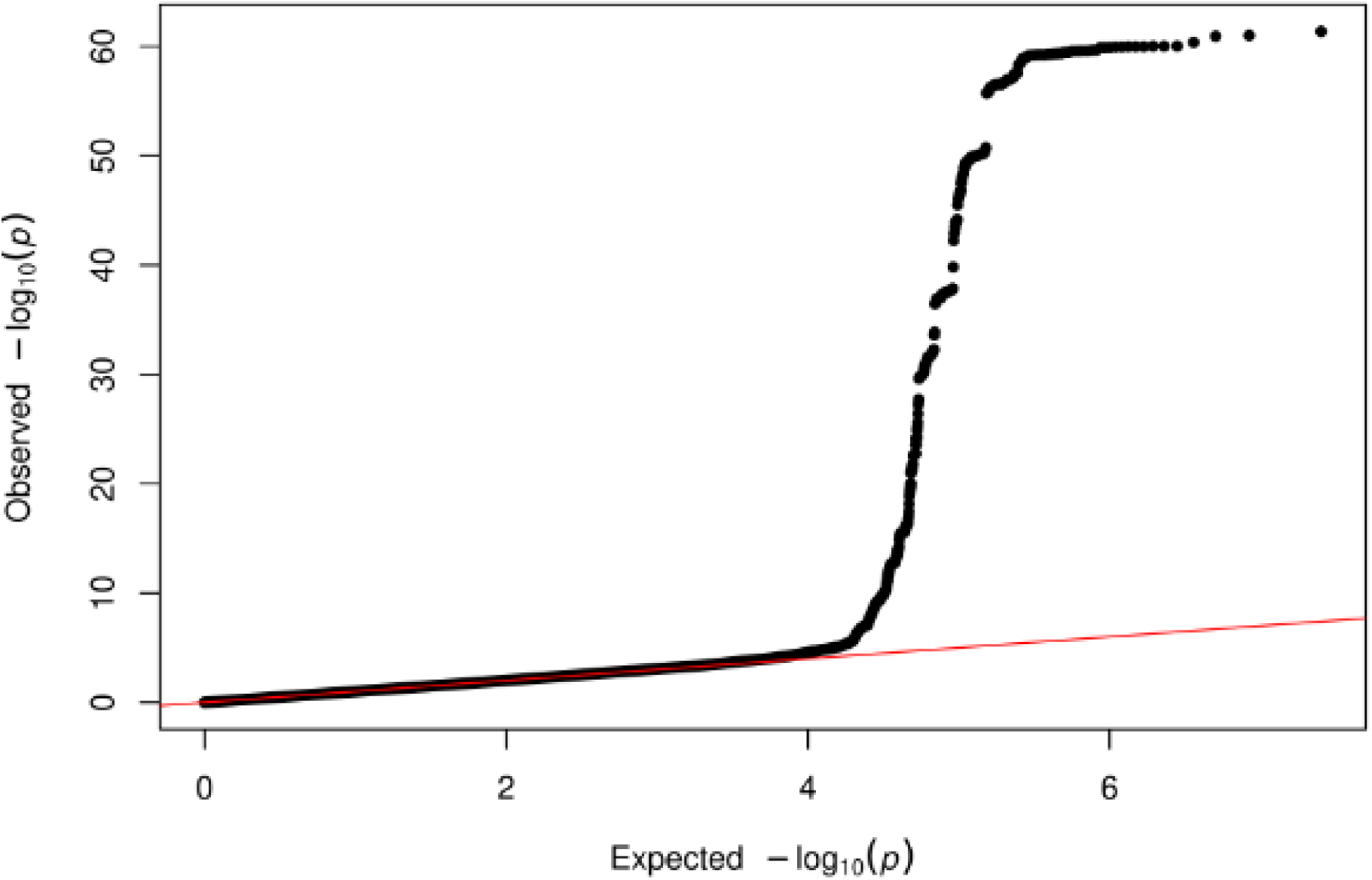
Quantile-quantile plot (Q-Q plot) of GWAS for CSF sTREM2 in European individuals (EUR-GWAS). Q-Q plot of EUR-GWAS: P values are two-sided raw P values estimated from a linear additive model. Y-axis represents observed –log10(p) and the x-axis depicts the expected –log10(p).

**Fig. S3.**
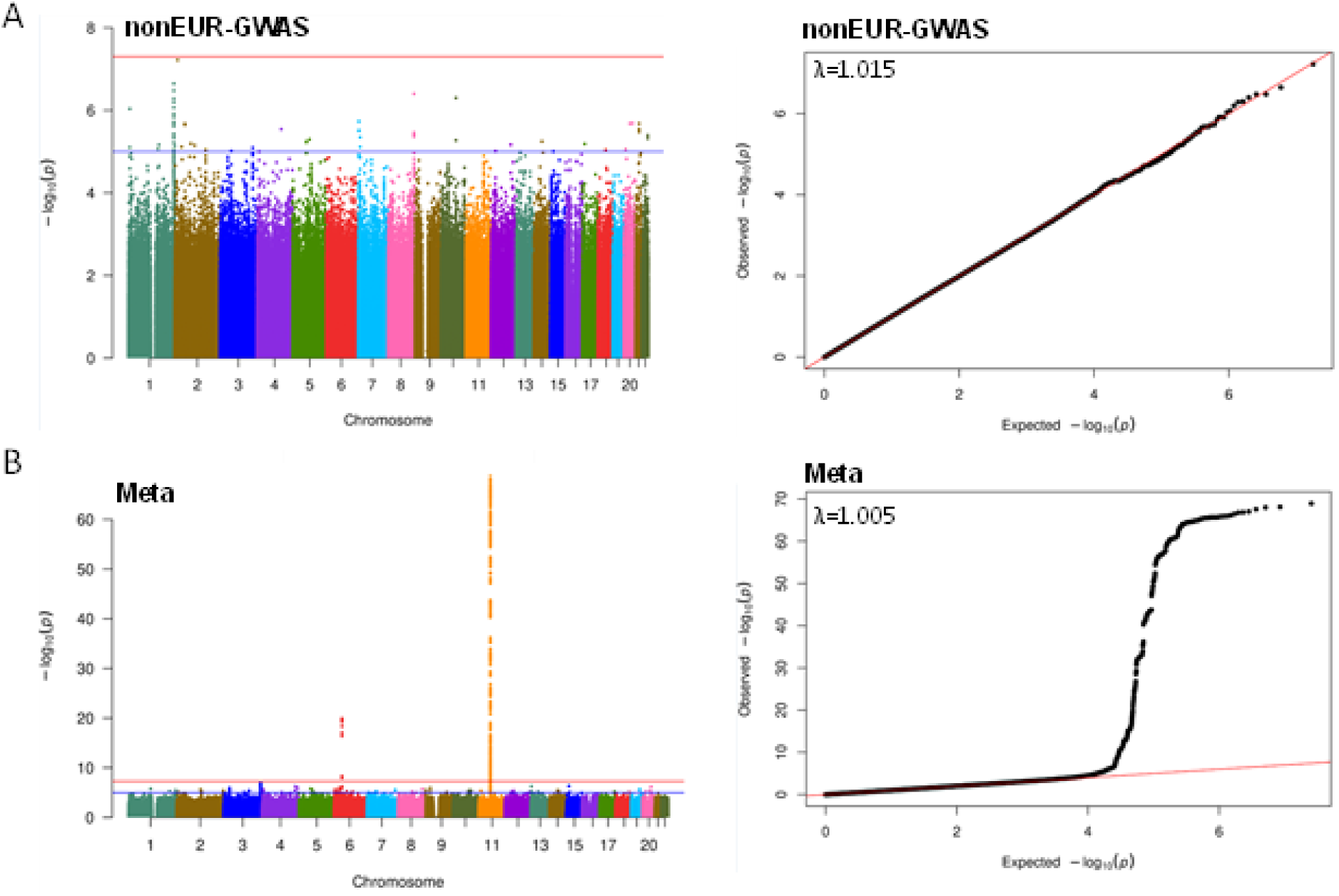
Manhattan plot and quantile-quantile plots (Q-Q plots) of GWAS for CSF sTREM2 in non-European individuals (nonEUR-GWAS) and meta-analyses of EUR GWAS and nonEUR GWAS. **A**) Left panel is Manhattan plots of nonEUR-GWAS. P values are two-sided raw P values estimated from a linear additive model. The blue solid horizontal line denotes the genome-wide significance level (P = 5 × 10-8), and the red solid horizontal line represents the suggestive significance level (P = 1 × 10-6). X-axis depicts genomic coordinates by chromosome number and y-axis denotes the negative log10-transformed P value for each genetic variant. Right panel is the Q-Q plot of nonEUR_GWAS. Y-axis represents observed –log10(p) and x-axis depicts the expected –log10(p). **B**) Left panel is Manhattan plots of meta-analyses of EUR-GWAS and nonEUR-GWAS for CSF sTREM2. P values are two-sided raw P values estimated from a linear additive model. The blue solid horizontal line denotes the genome-wide significance level (P = 5 × 10-8), and the red solid horizontal line represents the suggestive significance level (P = 1 × 10-6). X-axis depicts genomic coordinates by chromosome number and y-axis denotes the negative log10-transformed P value for each genetic variant. Right panel is the Q-Q plot of meta-analyses of EUR-GWAS and nonEUR-GWAS. Y-axis represents observed –log10(p) and x-axis depicts the expected –log10(p).

**Fig. S4.**
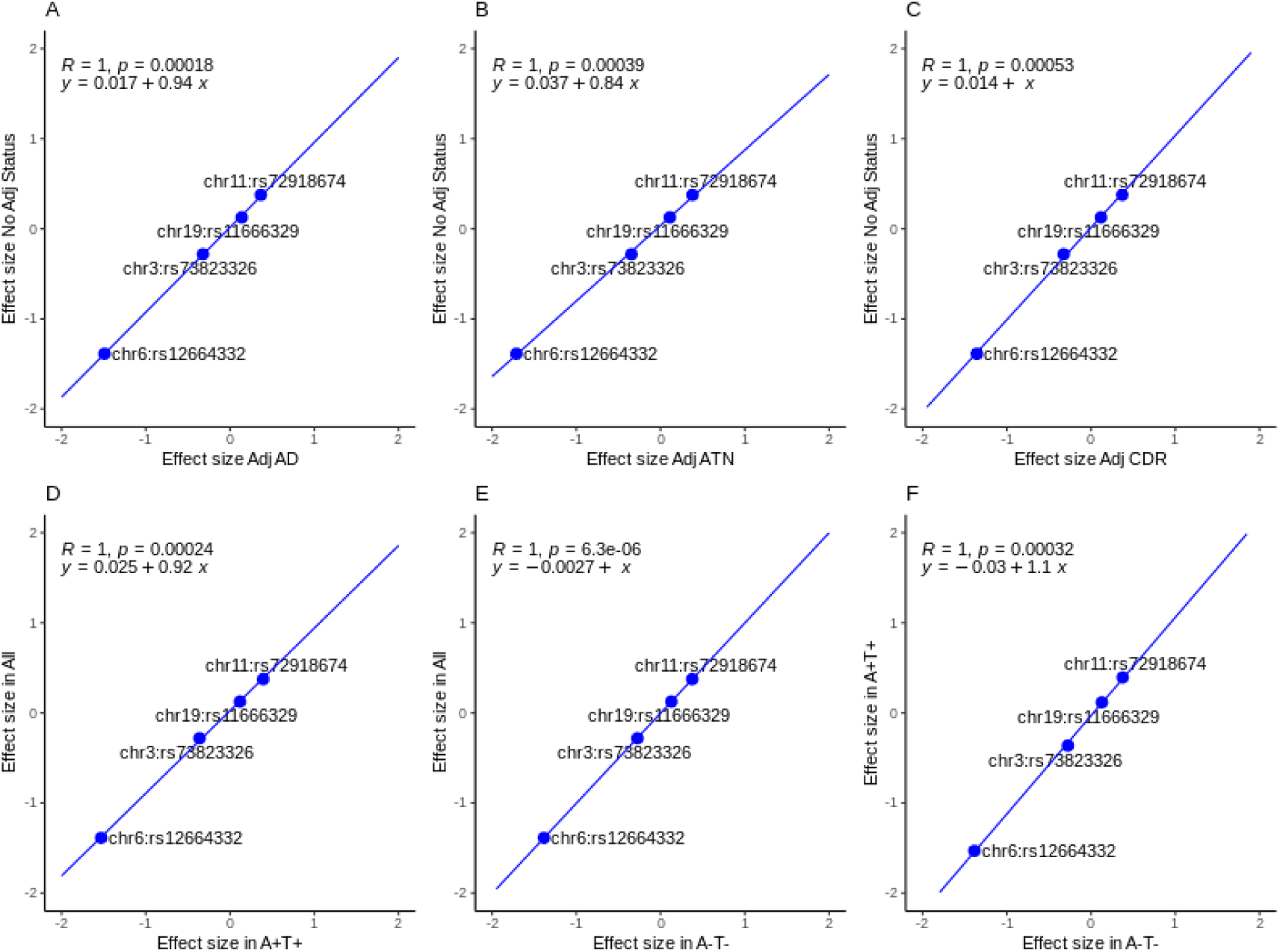
Scatterplot of effect size across GWAS analyses at four index variants. **A)** GWAS with and without adjusting for AD status. **B)** GWAS with and without AT classification. **C)** GWAS with and without adjusting for CDR. **D)** GWAS in all individuals vs. GWAS in biomarker positive (A+T+). **E)** GWAS in all vs GWAS in biomarker negative (A-T-). **F)** GWAS in biomarker positive (A+T+) vs. in biomarker negative (A-T). AD: Alzheimer’s disease; CDR: Clinical Dementia Rating.

**Fig. S5.**
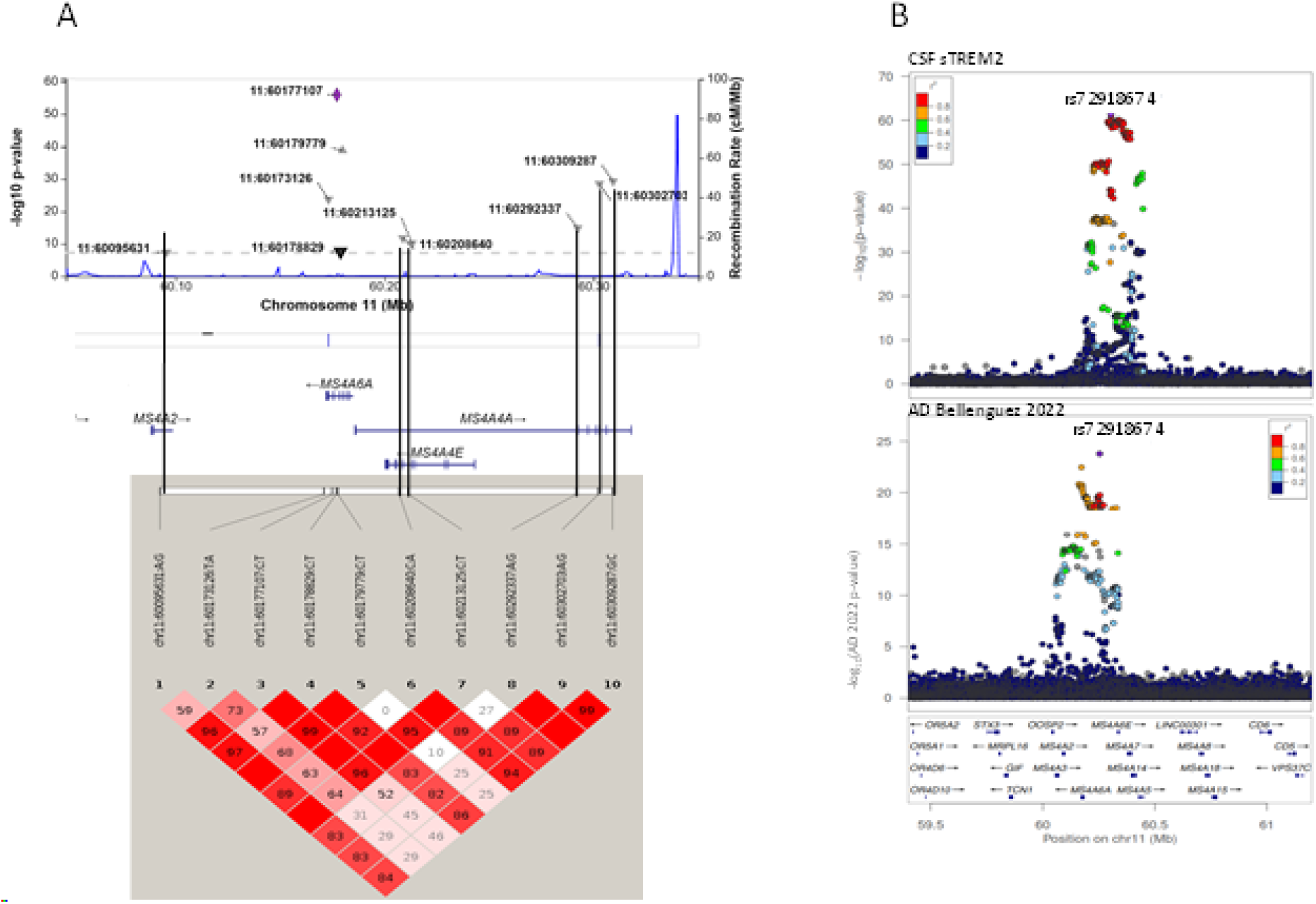
Association results of CSF sTREM2 at chromosome 11. **A)** LocusZoom plots at chromosome 11 for 10 SNPs (upper panel) and linkage disequilibrium (LD) heatmap of these 10 SNPs based on R2 (the square of the correlation coefficient) estimated using Haploview 4.2 (lower panel); **B)** LocusZoom plots at chromosome 11 for GWAS of CSF sTREM2 and GWAS of Alzheimer’s disease (AD) in 2022. X-axis depicts genomic coordinates at chromosome 11 and y-axis denotes the negative log10-transformed P value for each genetic variant.

**Fig. S6.**
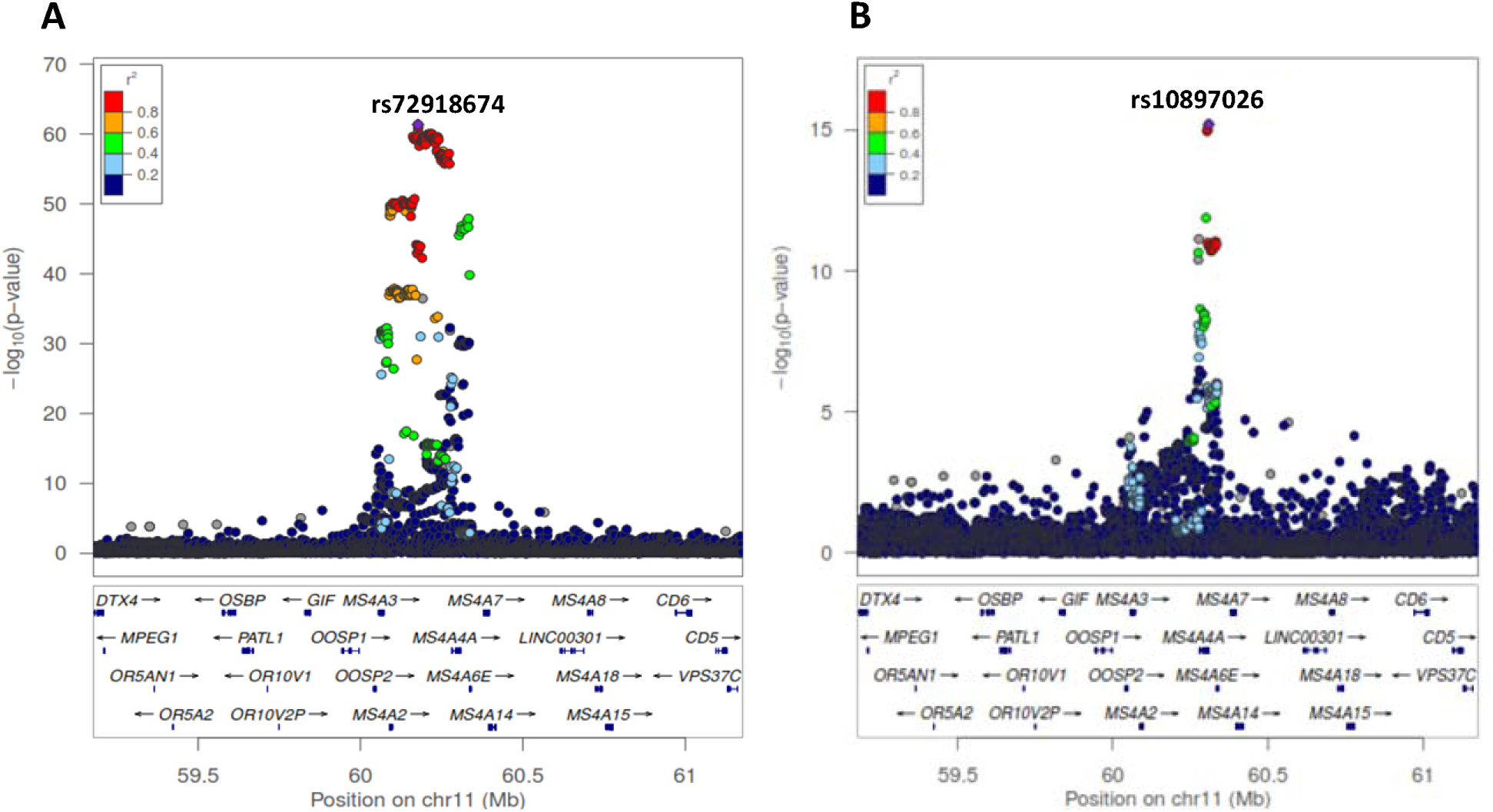
Locus plots of rs12664332 at chromosome 6 for CSF sTREM2 and AD. **A)** LocusZoom plots at chromosome 6 for GWAS of CSF sTREM2. B) LocusZoom plots at chromosome 6 for GWAS of AD in 2022. The X-axis depicts genomic coordinates at chromosome 11 and the y-axis denotes the negative log10-transformed P value for each genetic variant.

**Fig. S7.**
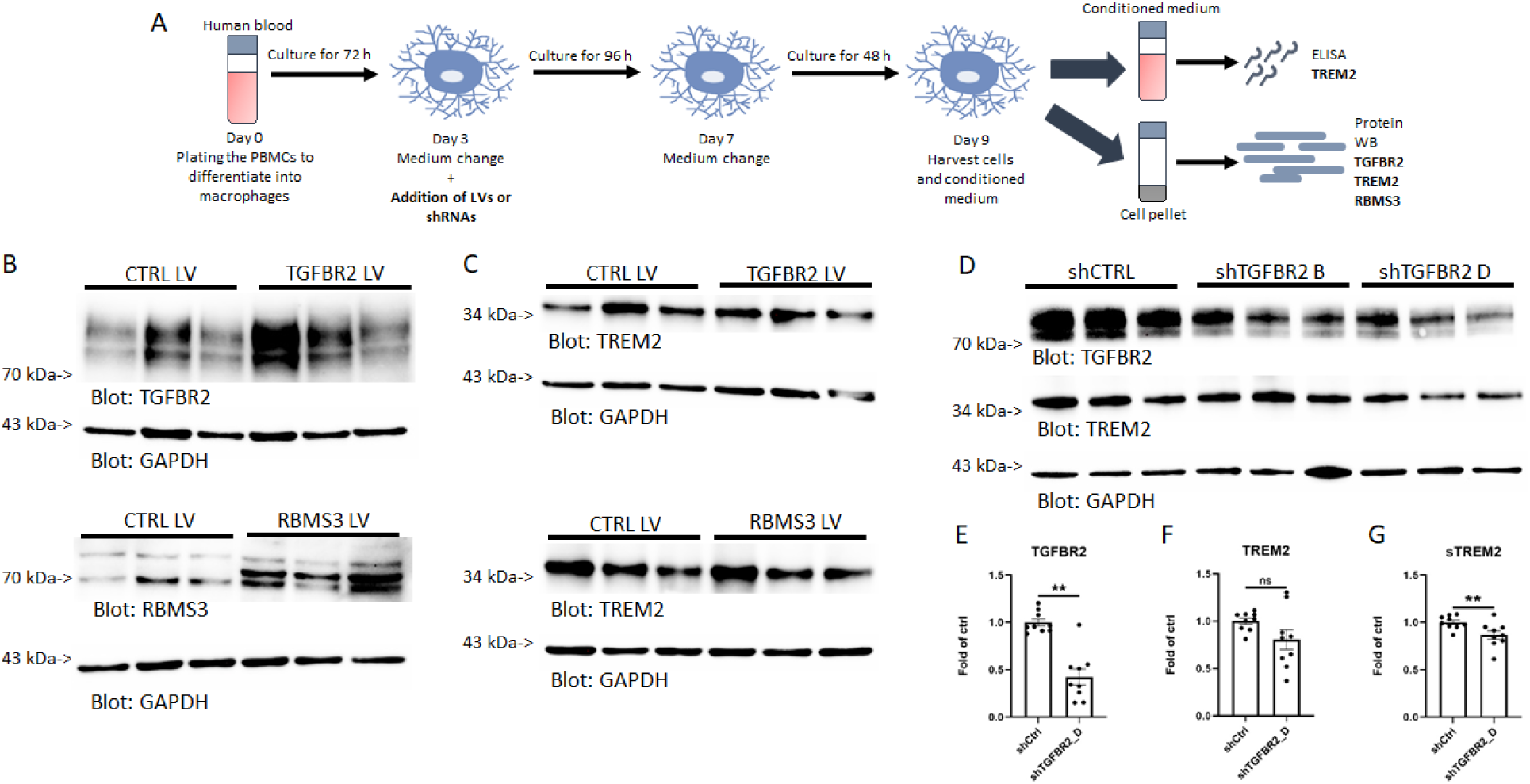
Experimental design, representative western blots of *TGFBR2* and *RBMS3* overexpression and knockdown of *TGFBR2* in PBMC-derived macrophages presented in Figure 4. **A)** Experimental design for PBMC-derived macrophages. **B)** Intracellular TGFBR2 and RBMS3 protein levels and **C)** TREM2 levels upon TGFBR2 and RBMS3 overexpression. **D)** Representative western blot upon TGFBR2 knock down. Quantification of intracellular **E)** TGFBR2, **F)** intracellular TREM2 and G) extracellular sTREM2 levels upon *TGFBR2* knockdown using TGFBR2_D shRNA. n = 9 from 3 independent experiments. ns: not significant, ** p < 0.01. Results are shown in mean ± SEM.

**Fig. S8.**
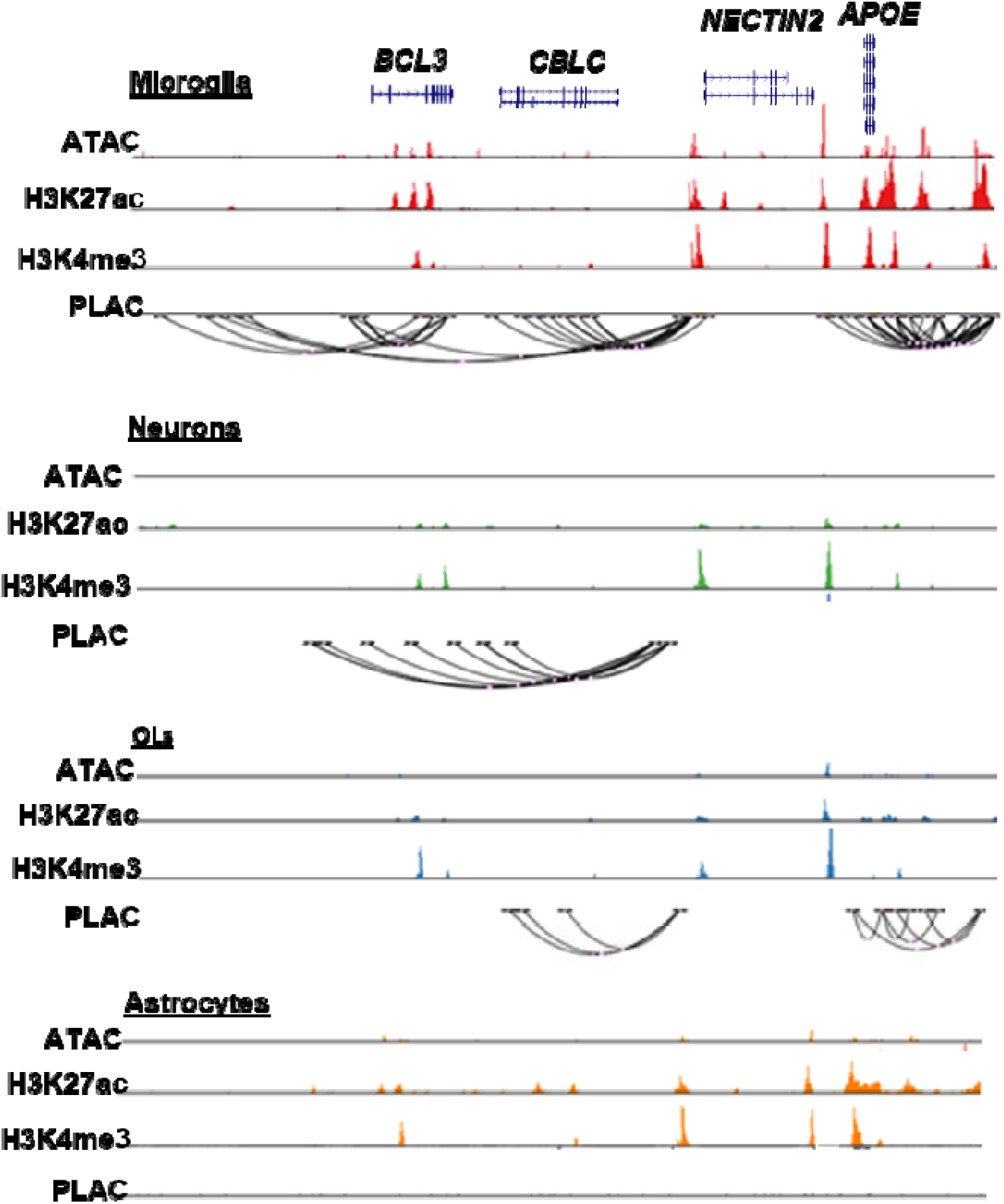
UCSC genome browser visualization of brain cell type specific ATAC-seq, H3K27ac ChiP-seq, H3K4me3 ChiP-seq and PLAC-seq loops at the chr 19 *APOE* locus. Chromatin loops link to promoters of *NECTIN2* to active gene-regulatory region are identified in microglia, neurons and oligodendrocytes (OLs).

**Fig. S9.**
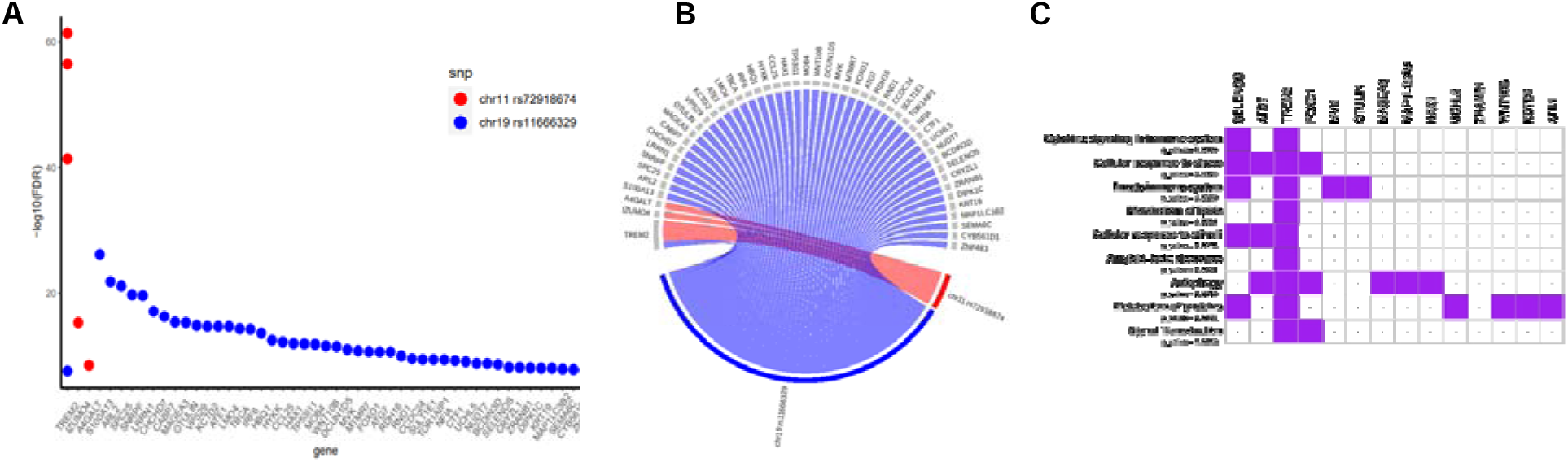
Dot plots and circular plots of proteins associated with rs72918674 at chromosome 11 and rs11666329 at chromosome 19. A) Dot plots of –log10(FDR) by proteins for rs72918674 and rs11666329. B) Circular plots of proteins associated with rs72918674 and rs11666329. C) Tile plots for pathways enriched in 47 proteins.

## Notes

### Author Declarations

The study was approved by the institutional review board of Washington University School of Medicine in St.Louis.

## References

1. Di Battista AM, Heinsinger NM, Rebeck GW: Alzheimer’s Disease Genetic Risk Factor APOE-epsilon4 Also Affects Normal Brain Function. Curr Alzheimer Res 2016, 13:1200–1207.

2. Jonsson T, Stefansson H, Steinberg S, Jonsdottir I, Jonsson PV, Snaedal J, Bjornsson S, Huttenlocher J, Levey AI, Lah JJ, et al: Variant of TREM2 associated with the risk of Alzheimer’s disease. N Engl J Med 2013, 368:107–116.

3. Guerreiro R, Wojtas A, Bras J, Carrasquillo M, Rogaeva E, Majounie E, Cruchaga C, Sassi C, Kauwe JS, Younkin S, et al: TREM2 variants in Alzheimer’s disease. N Engl J Med 2013, 368:117–127.

4. Benitez BA, Cooper B, Pastor P, Jin SC, Lorenzo E, Cervantes S, Cruchaga C: TREM2 is associated with the risk of Alzheimer’s disease in Spanish population. Neurobiol Aging 2013, 34:1711 e1715–1717.

5. Jin SC, Benitez BA, Karch CM, Cooper B, Skorupa T, Carrell D, Norton JB, Hsu S, Harari O, Cai Y, et al: Coding variants in TREM2 increase risk for Alzheimer’s disease. Hum Mol Genet 2014, 23:5838–5846.

6. Jin SC, Carrasquillo MM, Benitez BA, Skorupa T, Carrell D, Patel D, Lincoln S, Krishnan S, Kachadoorian M, Reitz C, et al: TREM2 is associated with increased risk for Alzheimer’s disease in African Americans. Mol Neurodegener 2015, 10:19.

7. Kunkle BW, Grenier-Boley B, Sims R, Bis JC, Damotte V, Naj AC, Boland A, Vronskaya M, van der Lee SJ, Amlie-Wolf A, et al: Genetic meta-analysis of diagnosed Alzheimer’s disease identifies new risk loci and implicates Abeta, tau, immunity and lipid processing. Nat Genet 2019, 51:414–430.

8. Bellenguez C, Kucukali F, Jansen IE, Kleineidam L, Moreno-Grau S, Amin N, Naj AC, Campos-Martin R, Grenier-Boley B, Andrade V, et al: New insights into the genetic etiology of Alzheimer’s disease and related dementias. Nat Genet 2022, 54:412–436.

9. Yang J, Fu Z, Zhang X, Xiong M, Meng L, Zhang Z: TREM2 ectodomain and its soluble form in Alzheimer’s disease. J Neuroinflammation 2020, 17:204.

10. Kim SM, Mun BR, Lee SJ, Joh Y, Lee HY, Ji KY, Choi HR, Lee EH, Kim EM, Jang JH, et al: TREM2 promotes Abeta phagocytosis by upregulating C/EBPalpha-dependent CD36 expression in microglia. Sci Rep 2017, 7:11118.

11. Hong S, Beja-Glasser VF, Nfonoyim BM, Frouin A, Li S, Ramakrishnan S, Merry KM, Shi Q, Rosenthal A, Barres BA, et al: Complement and microglia mediate early synapse loss in Alzheimer mouse models. Science 2016, 352:712–716.

12. Paolicelli RC, Bolasco G, Pagani F, Maggi L, Scianni M, Panzanelli P, Giustetto M, Ferreira TA, Guiducci E, Dumas L, et al: Synaptic pruning by microglia is necessary for normal brain development. Science 2011, 333:1456–1458.

13. Schafer DP, Lehrman EK, Kautzman AG, Koyama R, Mardinly AR, Yamasaki R, Ransohoff RM, Greenberg ME, Barres BA, Stevens B: Microglia sculpt postnatal neural circuits in an activity and complement-dependent manner. Neuron 2012, 74:691–705.

14. Wang Y, Cella M, Mallinson K, Ulrich JD, Young KL, Robinette ML, Gilfillan S, Krishnan GM, Sudhakar S, Zinselmeyer BH, et al: TREM2 lipid sensing sustains the microglial response in an Alzheimer’s disease model. Cell 2015, 160:1061–1071.

15. Lee CYD, Daggett A, Gu X, Jiang LL, Langfelder P, Li X, Wang N, Zhao Y, Park CS, Cooper Y, et al: Elevated TREM2 Gene Dosage Reprograms Microglia Responsivity and Ameliorates Pathological Phenotypes in Alzheimer’s Disease Models. Neuron 2018, 97:1032–1048 e1035.

16. Leyns CEG, Ulrich JD, Finn MB, Stewart FR, Koscal LJ, Remolina Serrano J, Robinson GO, Anderson E, Colonna M, Holtzman DM: TREM2 deficiency attenuates neuroinflammation and protects against neurodegeneration in a mouse model of tauopathy. Proc Natl Acad Sci U S A 2017, 114:11524–11529.

17. Jay TR, von Saucken VE, Landreth GE: TREM2 in Neurodegenerative Diseases. Mol Neurodegener 2017, 12:56.

18. Kiianitsa K, Kurtz I, Beeman N, Matsushita M, Chien WM, Raskind WH, Korvatska O: Novel TREM2 splicing isoform that lacks the V-set immunoglobulin domain is abundant in the human brain. J Leukoc Biol 2021, 110:829–837.

19. Filipello F, Goldsbury C, You SF, Locca A, Karch CM, Piccio L: Soluble TREM2: Innocent bystander or active player in neurological diseases? Neurobiol Dis 2022, 165:105630.

20. Kleinberger G, Yamanishi Y, Suarez-Calvet M, Czirr E, Lohmann E, Cuyvers E, Struyfs H, Pettkus N, Wenninger-Weinzierl A, Mazaheri F, et al: TREM2 mutations implicated in neurodegeneration impair cell surface transport and phagocytosis. Sci Transl Med 2014, 6:243ra286.

21. Piccio L, Deming Y, Del-Aguila JL, Ghezzi L, Holtzman DM, Fagan AM, Fenoglio C, Galimberti D, Borroni B, Cruchaga C: Cerebrospinal fluid soluble TREM2 is higher in Alzheimer disease and associated with mutation status. Acta Neuropathol 2016, 131:925–933.

22. Suarez-Calvet M, Kleinberger G, Araque Caballero MA, Brendel M, Rominger A, Alcolea D, Fortea J, Lleo A, Blesa R, Gispert JD, et al: sTREM2 cerebrospinal fluid levels are a potential biomarker for microglia activity in early-stage Alzheimer’s disease and associate with neuronal injury markers. EMBO Mol Med 2016, 8:466–476.

23. Suarez-Calvet M, Araque Caballero MA, Kleinberger G, Bateman RJ, Fagan AM, Morris JC, Levin J, Danek A, Ewers M, Haass C, Dominantly Inherited Alzheimer N: Early changes in CSF sTREM2 in dominantly inherited Alzheimer’s disease occur after amyloid deposition and neuronal injury. Sci Transl Med 2016, 8:369ra178.

24. Park SH, Lee EH, Kim HJ, Jo S, Lee S, Seo SW, Park HH, Koh SH, Lee JH: The relationship of soluble TREM2 to other biomarkers of sporadic Alzheimer’s disease. Sci Rep 2021, 11:13050.

25. Ewers M, Franzmeier N, Suarez-Calvet M, Morenas-Rodriguez E, Caballero MAA, Kleinberger G, Piccio L, Cruchaga C, Deming Y, Dichgans M, et al: Increased soluble TREM2 in cerebrospinal fluid is associated with reduced cognitive and clinical decline in Alzheimer’s disease. Sci Transl Med 2019, 11.

26. Ewers M, Biechele G, Suarez-Calvet M, Sacher C, Blume T, Morenas-Rodriguez E, Deming Y, Piccio L, Cruchaga C, Kleinberger G, et al: Higher CSF sTREM2 and microglia activation are associated with slower rates of beta-amyloid accumulation. EMBO Mol Med 2020, 12:e12308.

27. Deming Y, Filipello F, Cignarella F, Cantoni C, Hsu S, Mikesell R, Li Z, Del-Aguila JL, Dube U, Farias FG, et al: The MS4A gene cluster is a key modulator of soluble TREM2 and Alzheimer’s disease risk. Sci Transl Med 2019, 11.

28. Hou XH, Bi YL, Tan MS, Xu W, Li JQ, Shen XN, Dou KX, Tan CC, Tan L, Alzheimer’s Disease Neuroimaging I, Yu JT: Genome-wide association study identifies Alzheimer’s risk variant in MS4A6A influencing cerebrospinal fluid sTREM2 levels. Neurobiol Aging 2019, 84:241 e213–241 e220.

29. Gu S, Pakstis AJ, Li H, Speed WC, Kidd JR, Kidd KK: Significant variation in haplotype block structure but conservation in tagSNP patterns among global populations. Eur J Hum Genet 2007, 15:302–312.

30. Schwartzentruber J, Cooper S, Liu JZ, Barrio-Hernandez I, Bello E, Kumasaka N, Young AMH, Franklin RJM, Johnson T, Estrada K, et al: Genome-wide meta-analysis, fine-mapping and integrative prioritization implicate new Alzheimer’s disease risk genes. Nat Genet 2021, 53:392–402.

31. Jansen IE, van der Lee SJ, Gomez-Fonseca D, de Rojas I, Dalmasso MC, Grenier-Boley B, Zettergren A, Mishra A, Ali M, Andrade V, et al: Genome-wide meta-analysis for Alzheimer’s disease cerebrospinal fluid biomarkers. Acta Neuropathol 2022, 144:821–842.

32. Huang KL, Marcora E, Pimenova AA, Di Narzo AF, Kapoor M, Jin SC, Harari O, Bertelsen S, Fairfax BP, Czajkowski J, et al: A common haplotype lowers PU.1 expression in myeloid cells and delays onset of Alzheimer’s disease. Nat Neurosci 2017, 20:1052–1061.

33. Del-Aguila JL, Fernandez MV, Schindler S, Ibanez L, Deming Y, Ma S, Saef B, Black K, Budde J, Norton J, et al: Assessment of the Genetic Architecture of Alzheimer’s Disease Risk in Rate of Memory Decline. J Alzheimers Dis 2018, 62:745–756.

34. Nott A, Holtman IR, Coufal NG, Schlachetzki JCM, Yu M, Hu R, Han CZ, Pena M, Xiao J, Wu Y, et al: Brain cell type-specific enhancer-promoter interactome maps and disease-risk association. Science 2019, 366:1134–1139.

35. Vosa U, Claringbould A, Westra HJ, Bonder MJ, Deelen P, Zeng B, Kirsten H, Saha A, Kreuzhuber R, Yazar S, et al: Large-scale cis- and trans-eQTL analyses identify thousands of genetic loci and polygenic scores that regulate blood gene expression. Nat Genet 2021, 53:1300–1310.

36. Consortium GT: The GTEx Consortium atlas of genetic regulatory effects across human tissues. Science 2020, 369:1318–1330.

37. de Klein N, Tsai EA, Vochteloo M, Baird D, Huang Y, Chen CY, van Dam S, Oelen R, Deelen P, Bakker OB, et al: Brain expression quantitative trait locus and network analyses reveal downstream effects and putative drivers for brain-related diseases. Nat Genet 2023.

38. Lopes KP, Snijders GJL, Humphrey J, Allan A, Sneeboer MAM, Navarro E, Schilder BM, Vialle RA, Parks M, Missall R, et al: Genetic analysis of the human microglial transcriptome across brain regions, aging and disease pathologies. Nat Genet 2022, 54:4–17.

39. Phillips B, Western D, Wang L, Timsina J, Sun Y, Gorijala P, Yang C, Do A, Nykänen N-P, Alvarez I, et al: Proteome Wide Association Studies of LRRK2 variants identify novel causal and druggable for Parkinson’s disease. medRxiv 2023:2023.2001.2005.23284241.

40. Akiyama M, Ishigaki K, Sakaue S, Momozawa Y, Horikoshi M, Hirata M, Matsuda K, Ikegawa S, Takahashi A, Kanai M, et al: Characterizing rare and low-frequency height-associated variants in the Japanese population. Nat Commun 2019, 10:4393.

41. Barton AR, Sherman MA, Mukamel RE, Loh PR: Whole-exome imputation within UK Biobank powers rare coding variant association and fine-mapping analyses. Nat Genet 2021, 53:1260–1269.

42. Kichaev G, Bhatia G, Loh PR, Gazal S, Burch K, Freund MK, Schoech A, Pasaniuc B, Price AL: Leveraging Polygenic Functional Enrichment to Improve GWAS Power. Am J Hum Genet 2019, 104:65–75.

43. Vuckovic D, Bao EL, Akbari P, Lareau CA, Mousas A, Jiang T, Chen MH, Raffield LM, Tardaguila M, Huffman JE, et al: The Polygenic and Monogenic Basis of Blood Traits and Diseases. Cell 2020, 182:1214–1231 e1211.

44. Schumacher FR, Al Olama AA, Berndt SI, Benlloch S, Ahmed M, Saunders EJ, Dadaev T, Leongamornlert D, Anokian E, Cieza-Borrella C, et al: Association analyses of more than 140,000 men identify 63 new prostate cancer susceptibility loci. Nat Genet 2018, 50:928–936.

45. Chen MH, Raffield LM, Mousas A, Sakaue S, Huffman JE, Moscati A, Trivedi B, Jiang T, Akbari P, Vuckovic D, et al: Trans-ethnic and Ancestry-Specific Blood-Cell Genetics in 746,667 Individuals from 5 Global Populations. Cell 2020, 182:1198–1213 e1114.

46. Park JS, Ji IJ, Kim DH, An HJ, Yoon SY: The Alzheimer’s Disease-Associated R47H Variant of TREM2 Has an Altered Glycosylation Pattern and Protein Stability. Front Neurosci 2016, 10:618.

47. Suarez-Calvet M, Morenas-Rodriguez E, Kleinberger G, Schlepckow K, Araque Caballero MA, Franzmeier N, Capell A, Fellerer K, Nuscher B, Eren E, et al: Early increase of CSF sTREM2 in Alzheimer’s disease is associated with tau related-neurodegeneration but not with amyloid-beta pathology. Mol Neurodegener 2019, 14:1.

48. Tesseur I, Zou K, Esposito L, Bard F, Berber E, Can JV, Lin AH, Crews L, Tremblay P, Mathews P, et al: Deficiency in neuronal TGF-beta signaling promotes neurodegeneration and Alzheimer’s pathology. J Clin Invest 2006, 116:3060–3069.

49. Zoller T, Schneider A, Kleimeyer C, Masuda T, Potru PS, Pfeifer D, Blank T, Prinz M, Spittau B: Silencing of TGFbeta signalling in microglia results in impaired homeostasis. Nat Commun 2018, 9:4011.

50. Mizutani K, Miyata M, Shiotani H, Kameyama T, Takai Y: Nectin-2 in general and in the brain. Mol Cell Biochem 2022, 477:167–180.

51. Miyata M, Mandai K, Maruo T, Sato J, Shiotani H, Kaito A, Itoh Y, Wang S, Fujiwara T, Mizoguchi A, et al: Localization of nectin-2delta at perivascular astrocytic endfoot processes and degeneration of astrocytes and neurons in nectin-2 knockout mouse brain. Brain Res 2016, 1649:90–101.

52. Logue MW, Schu M, Vardarajan BN, Buros J, Green RC, Go RC, Griffith P, Obisesan TO, Shatz R, Borenstein A, et al: A comprehensive genetic association study of Alzheimer disease in African Americans. Arch Neurol 2011, 68:1569–1579.

53. Bailey CC, DeVaux LB, Farzan M: The Triggering Receptor Expressed on Myeloid Cells 2 Binds Apolipoprotein E. J Biol Chem 2015, 290:26033–26042.

54. Atagi Y, Liu CC, Painter MM, Chen XF, Verbeeck C, Zheng H, Li X, Rademakers R, Kang SS, Xu H, et al: Apolipoprotein E Is a Ligand for Triggering Receptor Expressed on Myeloid Cells 2 (TREM2). J Biol Chem 2015, 290:26043–26050.

55. Edwin TH, Henjum K, Nilsson LNG, Watne LO, Persson K, Eldholm RS, Saltvedt I, Halaas NB, Selbaek G, Engedal K, et al: A high cerebrospinal fluid soluble TREM2 level is associated with slow clinical progression of Alzheimer’s disease. Alzheimers Dement (Amst) 2020, 12:e12128.

56. Moreno-Grau S, de Rojas I, Hernandez I, Quintela I, Montrreal L, Alegret M, Hernandez-Olasagarre B, Madrid L, Gonzalez-Perez A, Maronas O, et al: Genome-wide association analysis of dementia and its clinical endophenotypes reveal novel loci associated with Alzheimer’s disease and three causality networks: The GR@ACE project. Alzheimers Dement 2019, 15:1333–1347.

57. Alvarez I, Diez-Fairen M, Aguilar M, Gonzalez JM, Ysamat M, Tartari JP, Carcel M, Alonso A, Brix B, Arendt P, Pastor P: Added value of cerebrospinal fluid multimarker analysis in diagnosis and progression of dementia. Eur J Neurol 2021, 28:1142–1152.

58. Marek K, Chowdhury S, Siderowf A, Lasch S, Coffey CS, Caspell-Garcia C, Simuni T, Jennings D, Tanner CM, Trojanowski JQ, et al: The Parkinson’s progression markers initiative (PPMI) - establishing a PD biomarker cohort. Ann Clin Transl Neurol 2018, 5:1460–1477.

59. Winfree RL, Dumitrescu L, Blennow K, Zetterberg H, Gifford KA, Pechman KR, Jefferson AL, Hohman TJ, Alzheimer’s Disease Neuroimaging I: Biological correlates of elevated soluble TREM2 in cerebrospinal fluid. Neurobiol Aging 2022, 118:88–98.

60. Gold L, Ayers D, Bertino J, Bock C, Bock A, Brody EN, Carter J, Dalby AB, Eaton BE, Fitzwater T, et al: Aptamer-based multiplexed proteomic technology for biomarker discovery. PLoS One 2010, 5:e15004.

61. Candia J, Cheung F, Kotliarov Y, Fantoni G, Sellers B, Griesman T, Huang J, Stuccio S, Zingone A, Ryan BM, et al: Assessment of Variability in the SOMAscan Assay. Sci Rep 2017, 7:14248.

62. Chang CC, Chow CC, Tellier LC, Vattikuti S, Purcell SM, Lee JJ: Second-generation PLINK: rising to the challenge of larger and richer datasets. Gigascience 2015, 4:7.

63. Willer CJ, Li Y, Abecasis GR: METAL: fast and efficient meta-analysis of genomewide association scans. Bioinformatics 2010, 26:2190–2191.

64. Pruim RJ, Welch RP, Sanna S, Teslovich TM, Chines PS, Gliedt TP, Boehnke M, Abecasis GR, Willer CJ: LocusZoom: regional visualization of genome-wide association scan results. Bioinformatics 2010, 26:2336–2337.

65. Barrett JC, Fry B, Maller J, Daly MJ: Haploview: analysis and visualization of LD and haplotype maps. Bioinformatics 2005, 21:263–265.

66. Yang J, Lee SH, Goddard ME, Visscher PM: GCTA: a tool for genome-wide complex trait analysis. Am J Hum Genet 2011, 88:76–82.

67. McLaren W, Gil L, Hunt SE, Riat HS, Ritchie GR, Thormann A, Flicek P, Cunningham F: The Ensembl Variant Effect Predictor. Genome Biol 2016, 17:122.

68. Hemani G, Zheng J, Elsworth B, Wade KH, Haberland V, Baird D, Laurin C, Burgess S, Bowden J, Langdon R, et al: The MR-Base platform supports systematic causal inference across the human phenome. Elife 2018, 7.

69. Giambartolomei C, Vukcevic D, Schadt EE, Franke L, Hingorani AD, Wallace C, Plagnol V: Bayesian test for colocalisation between pairs of genetic association studies using summary statistics. PLoS Genet 2014, 10:e1004383.

70. Choi SW, O’Reilly PF: PRSice-2: Polygenic Risk Score software for biobank-scale data. Gigascience 2019, 8.

71. Ferkingstad E, Sulem P, Atlason BA, Sveinbjornsson G, Magnusson MI, Styrmisdottir EL, Gunnarsdottir K, Helgason A, Oddsson A, Halldorsson BV, et al: Large-scale integration of the plasma proteome with genetics and disease. Nat Genet 2021, 53:1712–1721.

